# A Novel Method to Disentangle Tightly Linked Risk and Resilience Genes for Brain Disorders: Application to Alzheimer’s Disease

**DOI:** 10.1101/2025.02.26.25322962

**Authors:** Eric J Barnett, Jonathan L Hess, Jiahui Hou, Valentina Escott-Price, Christine Fennema-Notestine, William Kremen, Shu-Ju Lin, Chunling Zhang, Chris Gaiteri, Jeremy Elman, Peter Holmans, Stephen V Faraone, Stephen J Glatt

## Abstract

Genetic risk factors for neuropsychiatric disorders are well documented. However, some individuals with high genetic risk remain unaffected, and the mechanisms underlying such resilience remain poorly understood. The presence of protective resilience factors that mitigate risk could help explain the disconnect between predicted risk and reality, particularly when genetic contributions are substantial but incompletely understood. Identifying and studying resilience factors could improve our understanding of pathology, enhance risk prediction, and inform preventive measures or treatment strategies. However, such efforts are complicated by the difficulty of identifying resilience that is separable from low risk. We developed a novel adversarial multi-task neural network model to detect genetic resilience markers. The model learns to separate high-risk unaffected individuals from affected individuals at similar risk while “unlearning” patterns found in low-risk groups using adversarial learning. In simulated and existing Alzheimer’s disease (AD) datasets, we identified markers of resilience with a feature-importance-based approach that prioritized specificity, generated resilience scores, and analyzed associations with polygenic risk scores (PRS). In simulations, our model had high specificity and sensitivity in identifying resilience markers, significantly outperforming traditional approaches. Applied to AD data, the model generated genetic resilience scores protective against AD and independent of PRS. We identified five resilience-associated SNPs, including known AD-associated variants, underscoring their potential involvement in resilience. Our findings support the utility of resilience scores in modifying risk predictions, particularly for high-risk groups. Expanding this method could aid in understanding resilience mechanisms, potentially improving diagnosis, prevention, and treatment strategies for AD and other brain disorders.

## Introduction

Through global research efforts, great strides have been made in improving our understanding of the genetic risk factors associated with psychiatric and neurodegenerative disorders. Despite all we have learned about risk factors for these disorders, it is still unclear as to why some people with high risk for a disorder remain unaffected. One explanation is that those people have protective features that make them resilient to developing the disorder. Resilience in this context refers to features that mitigate the impact of risk factors, or in the prevailing taxonomy, these factors increase resilient “capacity”.(1, 2) A better understanding of resilience would improve our ability to predict risk for psychiatric and neurodegenerative disorders, leading to earlier and more objective interventions. Just as importantly, such knowledge would lend insight into avenues for better treatment and prevention.

The genetic architecture of Alzheimer’s disease (AD) indicates that resilience is a powerful lens through which to view pathogenesis and therapeutic options. Twin studies have estimated the heritability of late-onset Alzheimer’s disease (LOAD), the most common form of AD, at 58 – 79%,(3) but the variance explained by common single nucleotide polymorphisms (SNPs) in additive models is lower, at 38-66%, and has been found to be decreasing with larger study sizes.(4) Further understanding of AD will be critical in explaining this “missing heritability” and combating the disorder’s growing impacts on individuals and society. Indeed, studies have found that people with AD brain pathology are not destined to have or develop dementia. Both environmental and genetic resilience factors have been shown to play an important role in the pathology of AD.(5) In this study we focus on genetic resilience factors, which are sequence variants that modify the risk that would otherwise be associated with one or more loci.

Since the discovery of the risk-conferring *APOE* haplotypes, other studies have identified SNPs in nearby genes that modify the effects of *APOE,*(6, 7) which otherwise carries the greatest risk for LOAD of any gene. The *APOE* haplotype that confers AD risk is made up of two SNPs, rs429358 and rs7412.(8) When both SNPs are cytosine, as in the *APOE*-ε4 haplotype, the odds of developing AD are 3.7 times higher compared to the most common scenario where rs429358 is a thymine and rs7412 is a cytosine,(9) as is the case in the *APOE*-ε3 haplotype. When both SNPs are thymine, as in the *APOE*-ε2 haplotype, the odds ratio for developing AD is 0.6 compared to *APOE*-ε3, indicating a protective effect. The alleles at these two SNPs regulate *APOE* protein isoforms, which go on to affect the clearance, transport, and immune response pathways implicated in AD.(9) It could be inferred that the protein structure produced based on thymine alleles in rs429358 are protective against either the structural effects of the rs7412(C) allele or the combined effects of rs7412(C) and rs429358(C).(10) Many studies have nominated cis or trans elements that modify APOE risk.(11, 12) (7, 13–16) Such APOE results illustrate the broader challenge in which many genomic resilience features are positionally nearby and in LD with risk variants. Therefore, it would be helpful to develop a method that can include correlated variants while adjusting out pure risk effects.

Here, we introduce a novel neural network approach to detect markers of resilience with increased resolution. Specifically, it overcomes the requirement of prior methods to exclude SNPs in LD with risk SNPs, addressing a major limitation of previous approaches(17). We used multi-task neural networks to find markers of resilience, which we define as SNPs that accurately discriminate unaffected people who have high risk for a disorder from affected people with similar risk, but which cannot accurately discriminate low-risk unaffected from affected people with similar risk. In other words, genetic markers that collectively make high-risk individuals resilient to developing disease but have little to no effect on disease development among individuals at low risk. This approach was designed to simulate the real-world possibility of syntenic risk and resilience SNPs, and to test our ability to detect such resilience SNPs when they are in strong LD with risk SNPs. We hypothesized that the difference in allele frequency of these resilience SNPs would be higher when comparing high-risk unaffected and high-risk affected individuals relative to comparisons between low-risk unaffected and low-risk affected individuals. This is because, without resilience features, the elevated risk found in high risk-unaffected people would otherwise result in more people in this group developing the disorder, whereas the presence of resilience genes in individuals at low risk does not impact the likelihood of illness. We evaluated the validity of our approach by applying it to both simulated and previously collected AD genome-wide association study (GWAS) data.

## Methods and Materials

### Model Architecture

We designed a neural network architecture with *PyTorch*(18) to detect markers of resilience. We accomplished this using multi-task modeling. The first task of our model was to minimize error when predicting case vs. control status in the high-risk subgroup, which we defined as controls with a log-additive polygenic risk score (PRS) above a selected percentile in the control group distribution and PRS-matched cases. The second task was an adversarial task that *maximized* the error when predicting case vs. control status in the low-risk subgroup, which we defined as controls within a selected lower percentile range of the control group distribution and PRS-matched cases(19). This adversarial task differs from generative adversarial networks, which feature generator-discriminator pairs, and instead uses the domain-adversarial neural network approach to drive the model to unlearn any features that would be useful in predicting the low-risk subgroup. The PRS percentiles we used to define high-risk and low-risk subgroups were selected through a process we describe in the high-risk/low-risk subgroup thresholding analysis section. Unaccounted-for risk variants (i.e., those not identified in a typical comparison of all cases vs. all controls), and null variants are as likely to be present in the low-risk subgroup as they are to be in the high-risk subgroup, which means the adversarial task drives the model away from detecting and using those types of variants that might otherwise lead to false-positive resilience associations. The adversarial task was implemented by using a gradient reversal layer, which reverses the direction of the gradient during backpropagation such that the weights of the neural network are shifted in the direction that maximizes error for that task(19).

The neural network architecture is illustrated in Panel 6 of Figure 1. The primary and adversarial task in our models both have unique output layers but otherwise share the rest of the layers in the neural network. The gradient reversal layer is applied between the final shared layer and the adversarial task output layer, which means the adversarial task output layer is trying to minimize error in the task and will use any information available in the shared layers to best learn the adversarial task. With this design, we can accurately measure and optimize the difference in predictability between the high-risk subgroup and low-risk subgroup. For comparison, we also trained and optimized models with similar architectures but without the adversarial task. To maximize the difference between predictability in high-risk and low-risk subgroups, we optimized the learning rate, epochs, number of shared layers, shared layer size, L1 lambda, and the strength of the gradient reversal. The ranges for each of these hyperparameter optimizations can be found in Supplementary Table 1. Optimization was performed using *Optuna*(20) with a criterion of maximizing AUC for predicting case-control status in the high-risk subgroup and minimizing the difference between 0.5 and the AUC for predicting case-control status in the low-risk subgroup. Driving the AUC in the low-risk subgroup with the adversarial task is critical because it discourages the entire model from using any discriminative factors that are present in the low-risk subgroup. We anticipate that risk factors are likely to be similarly discriminative in high and low-risk subgroups. We expect that resilience factors and their interactions with risk factors are more discriminative in the high-risk subgroup, since those factors are likely to be more prevalent in individuals that remain unaffected despite high genetic risk. Therefore, the model is designed to model resilience factors and their interactions with risk factors in a way that is independent of log-additive risk.

**Figure 1:**
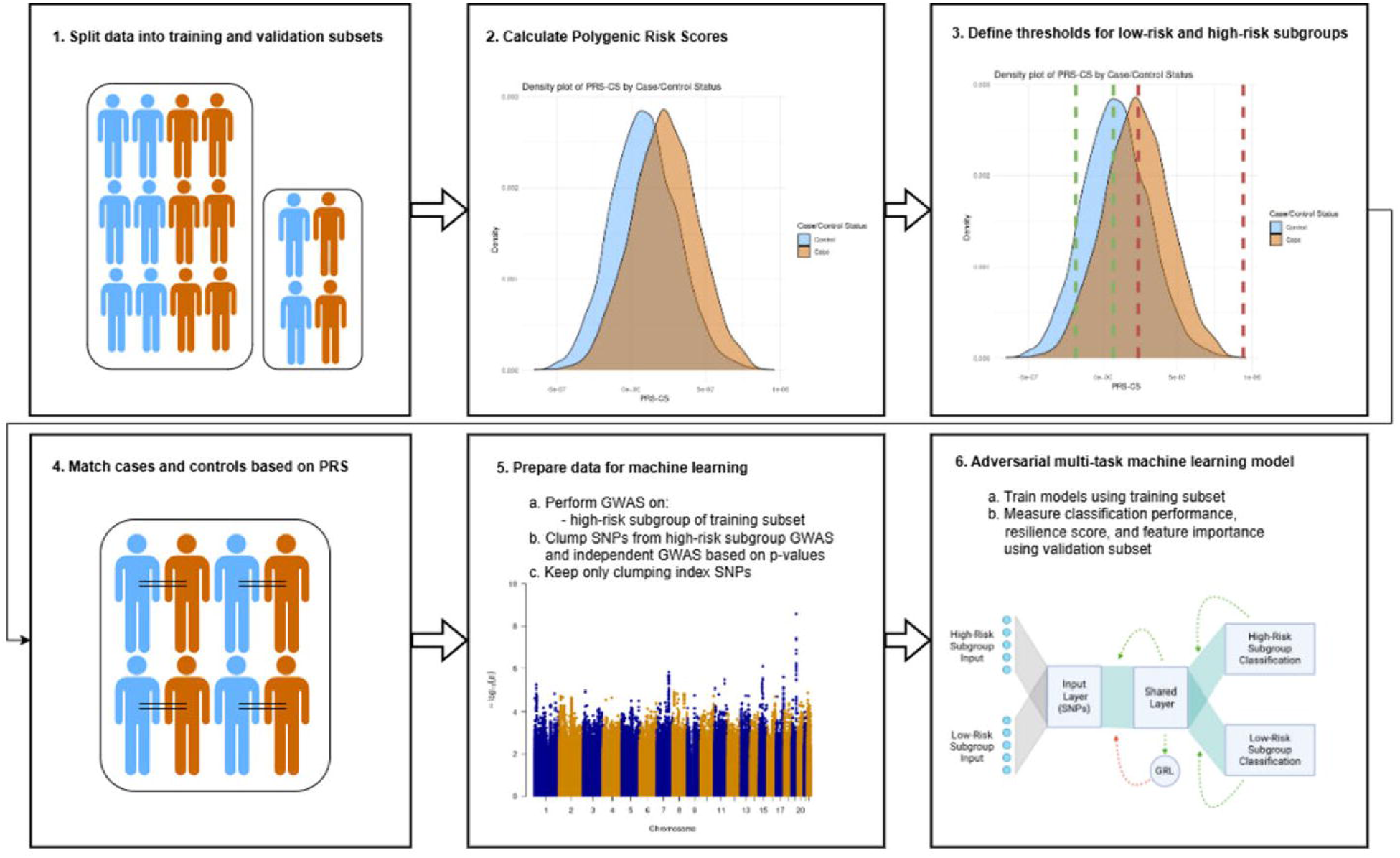
Alzheimer’s Disease Resilience Analysis Pipeline. Panel 1: We split data into a training subset (70%) used for GWAS and machine learning model training and validation (30%) used for reporting performance and feature importance. Panel 2: We calculate polygenic risk scores using independent, external summary statistics. Panel 3: We define minimum and maximum thresholds for low and high-risk subgroups and split the data based on those definitions. Panel 4: We match cases and controls based on PRS. Panel 5: We perform a GWAS on the high-risk subgroup in the training subset, clump SNPs from that GWAS and the external summary statistics based on AD association p-values and output the clumping index SNPs for machine learning modeling. Panel 6: We train models using the training subset in the illustrated model architecture. High-risk and low-risk subgroups are used simultaneously to collectively train the shared layer and used separately to train the risk-group-specific output layers. Backpropagation is modified for the low-risk subgroup classification task, such that after minimizing the error of the output layer, the gradient is reversed in the gradient reverse layer (GRL), resulting in an adversarial task that directs the model to maximize error for the low-risk subgroup task in the shared layer. In combination, the multi-task network directs the model to find and use features that are more predictive in the high-risk subgroup. We expect this group of features to be enriched with resilience features and their interactions with risk features. After model training, we measure classification performance and calculate resilience scores and feature importances using the validation subset.

### High-risk/Low-risk Subgroup Thresholding Analysis

Previous studies of genetic resilience have defined the resilient controls as those in the top 10% PRS in controls.(17, 21) For our machine learning models, we needed to define both a high-risk subgroup, where we expect to see resilience features more often in controls, and a low-risk subgroup, where we expect resilience features to be more evenly distributed. Ideally, we would aim to select subgroups such that we have high sensitivity for detecting markers of resilience and high specificity such that we will not falsely claim a SNP to be a marker of resilience when it is not. This would be a challenging task to optimize in real-world data since we cannot know for certain which SNPs are resilience markers. However, in our simulated data set we are able to set different thresholds and look at metrics for how well the models trained on the resulting subgroups specifically detect resilience SNPs. The methods we used to create the simulated data set can be found in the Supplementary Methods. We performed a grid search over a range of thresholds for defining the lower and upper thresholds for the low-risk subgroup and the lower threshold for the high-risk subgroup based on the PRS. The optimization ranges can be found in Supplementary Table 2. With each set of thresholds, we matched controls with cases and split the resulting data into a training subset containing 70% of the data to be used to train the models, a validation subset containing 15% of the data to be used to optimize the hyperparameters of the models, and a testing subset containing 15% of the data to be used for final measurement of performance metrics. We optimized models for each pair of high-risk and low-risk subgroups from each threshold possibility in our grid search. In hyperparameter optimization, we selected the model by finding the maximum value for the objective function

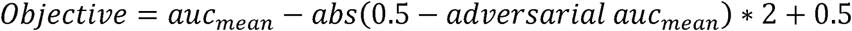

where auc_mean_ represents the mean AUC in the primary task across 5 model trials and adversarial auc_mean_ represents the mean AUC in the adversarial task across 5 model trials. This hyperparameter objective function effectively encourages models that are predictive in the primary task and have AUCs close to 0.5 in the adversarial task, with emphasis on keeping the adversarial task close to 0.5.

For the best model for each set of thresholds, we used *Integrated Gradients*(22) to identify the features that were most important in separating cases from controls. The output from *Integrated Gradients* is a feature-attribution value for each input feature for each simulated individual representing how much that feature contributed to the prediction of that simulated individual’s case/control status. To generate a more robust feature-attribution measurement, we performed *Integrated Gradients* analysis on 5 separately initialized and trained models and used the average attribution. To estimate overall feature importance, we compared the mean attributions in cases and controls for each feature using two-sided *t*-tests. We adjusted for multiple comparisons in our feature-importance analyses using Bonferroni correction.

Since our aim is to identify resilience features, we measured the sensitivity and specificity of our feature-importance analysis in identifying resilience SNPs while avoiding identifying other types of SNPs. For sensitivity and specificity calculations, we defined true positives as resilience SNPs that had significantly different attributions between cases and controls in the high-risk subgroup, false-positives as any other type of SNP that had significantly different attributions between cases and controls in the high-risk subgroup, false-negatives as resilience SNPs that did not have significantly different attributions in the high-risk subgroup, and true-negatives as any other type of SNP that did not have significantly different attributions in the high-risk subgroup. We also optimized models without the adversarial task for the same sets of thresholds and compared sensitivity and specificity across model type with paired t-tests.

For all models, we measured and reported AUC for predicting case-control status in the high-risk subgroup, AUC for predicting case-control status in the low-risk subgroup, and sensitivity and specificity in feature-importance analyses. While our models were not designed to maximize prediction, we calculated AUCs to compare the models’ prediction between the high-risk subgroup and low-risk subgroup and validate the models’ ability to find features that are more predictive in the high-risk subgroup.

### Alzheimer’s Disease Resilience Analysis

Our analysis pipeline is shown in Figure 1. We used the seven sets of Alzheimer’s Disease Center (ADC) genotyped subjects used in the Alzheimer’s Disease Genetics Consortium’s GWAS(23) to identify genes associated with an increased risk of developing AD. We applied the same sample and genotype quality-control steps as previous resilience analyses.(17) We randomly split the data into training (70%) and validation (30%) subsets. In the training subset, we used *PRS-CS*(24) with summary statistics from the Psychiatric Genomics Consortium’s AD GWAS,(25) which does not contain the data used in our analysis, and the 1000 Genomes Project reference panel(26) to infer posterior effects of SNPs after adjustment for LD. We used these posterior effects to create polygenic risk scores in the training and validation subsets. We split each subset into high-risk and low-risk subgroups based on these polygenic risk scores, using the set of definition thresholds that had the best balance of sensitivity and specificity in our simulation analyses. We then matched the controls in the high-risk subgroup with cases that had similar risk scores using the *MatchIt R*(27) package with the optimal pair matching method. We used the Welch two-sample *t*-test to calculate whether age was significantly different in cases and controls in the low-risk and high-risk subgroups. Likewise, we used Pearson’s chi-squared test to calculate whether sex or the presence of *APOE*-ε4 haplotypes were significantly different in cases and controls in the two subgroups.

In the matched training subset, we reduced the dimensions of the SNPs for input into our neural network models by clumping SNPs using the two *plink*(28) *clump* commands with default parameter settings. The first clumping command generated risk clumps by clumping based on SNP *p*-values from the Psychiatric Genomics Consortium AD GWAS. The second clumping command generated resilience clumps by clumping based on a GWAS of AD cases vs. controls performed in the matched, high-risk subgroup of the training subset. We also performed a GWAS in the matched, high-risk subgroup of the validation subset to compare to our neural network feature-importance results, but these test subset GWAS results were not used in any models. Resilience and traditional risk SNPs may be present in either group, but our rationale in using these two sets of SNPs is that our models need to have information about the risk for each person to look for interactions that resilience SNPs may have on that risk. For all subsets, we output a file containing the number of minor alleles for each SNP in the two SNP sets and the case/control label for each person. These files were the input for our neural network models.

We optimized hyperparameters using the same optimization ranges and objective function used in our simulation analysis and added dropout for additional regularization. Using the best set of hyperparameters, we performed the same *Integrated Gradients* feature-importance analysis we used in our simulation analysis. We used the average predictions across ten models for all of our data to generate a resilience score, which we defined as 1 minus the average prediction. This score reflected the tendency of neural networks to classify each individual as a control in our class-balanced, risk-matched, adversarial model. In this constrained model, we expect individuals predicted to be controls to be enriched in resilience features specific to the high-risk subgroup. We investigated the relationship between PRS and resilience score by removing any correlation between PRS-CS and resilience score through residualization and fitting a logistic regression in the training data predicting case/control status using PRS-CS, residualized resilience score, and the interaction between the PRS-CS and residualized resilience score.

We performed an additional analysis using the same methods restricted to individuals that are *APOE*-ε4 carriers. For this analysis, we removed *APOE* and its flanking region (chr19: 44,400 kb – 46,500 kb) from all steps. Hyperparameter optimization for this analysis failed to produce a reproducible model, so we did not perform further analysis on the resulting model.

## Results

### Simulated Data Analyses

Our SNP-pair simulation method created a sample of one million simulated individuals, all with 107 risk/null pairs, 149 inverse-risk/null pairs, 163 risk/resilience pairs, and 779 null/null pairs. The simulated data, each simulated individual’s standard additive risk and total risk scores, and information on each simulated SNP are available in the supplementary data file. Supplementary Figures 1 and 2 show density plots of the distribution of both risk scores in cases and controls.

**Figure 2:**
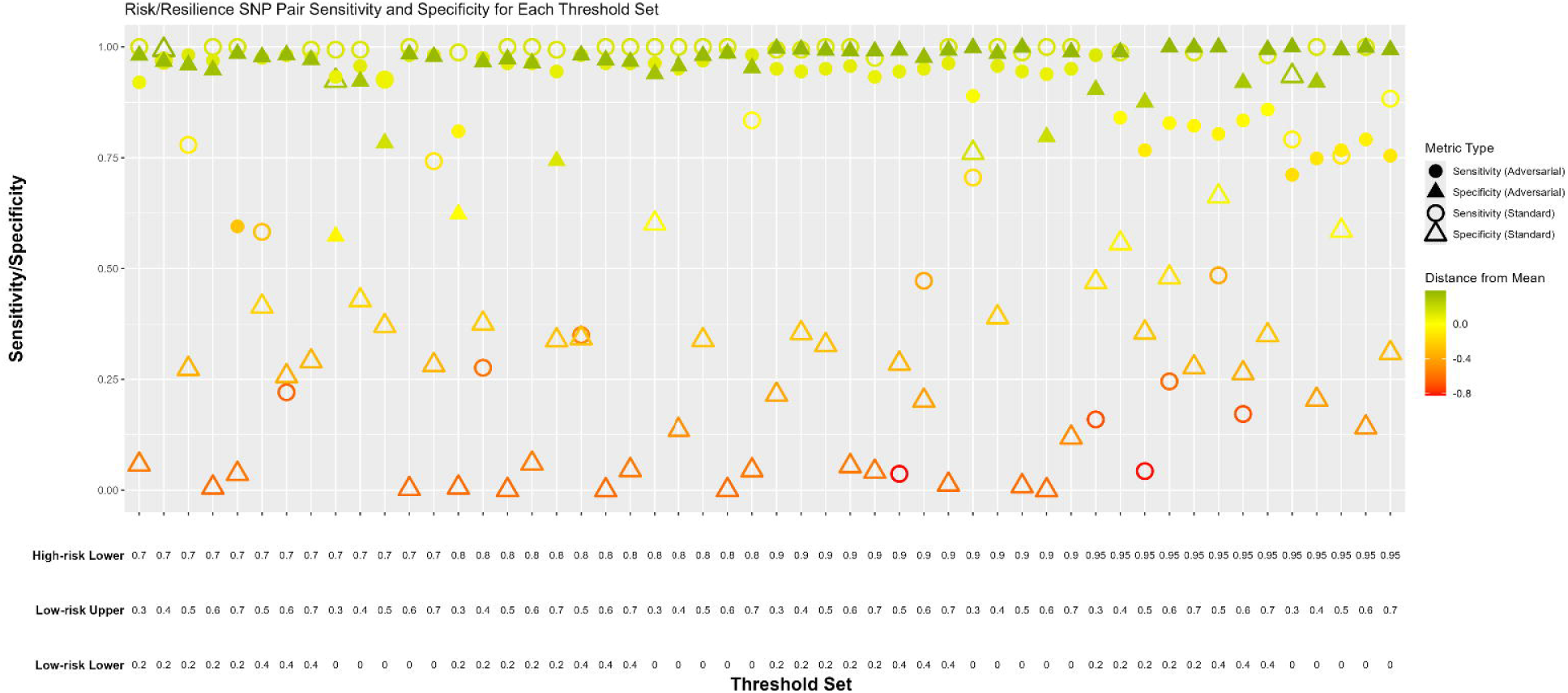
Identification of Resilience SNPs: Sensitivity and Specificity for each Threshold Set. Shown are the sensitivity and specificity in correctly detecting resilience SNPs across the tested threshold sets, which determined the PRS percentiles we used to define high-risk and low-risk subgroups. The threshold sets each contain a lower PRS percentile threshold for defining the high-risk subgroup, and a lower and upper PRS percentile threshold for defining the low-risk subgroup. Results are presented for models that had the adversarial task (filled shapes) and standard models that did not have the adversarial task (unfilled shapes) but were otherwise the same and were equally optimized. Our first priority in selecting the best model was high specificity, due to our goal of avoiding false positive results, and our second priority was high sensitivity. Models with adversarial tasks were on average significantly more specific and sensitive in identifying risk/resilience SNP-pairs.

The results of our thresholding analysis are shown in Figure 2 and Supplementary Table 2. Across all thresholds, when identifying resilience SNPs, the models with an adversarial task had significantly higher specificity (mean difference: 0.66, *p*-value < 2.2 × 10^-16^) and significantly higher sensitivity (mean difference: 0.10, *p*-value: 0.03) compared to models without an adversarial task. Based on the results, we chose to use the adversarial model using a threshold of 0.90 for the high-risk subgroup and thresholds of 0.2 and 0.5 for the low-risk subgroup because these thresholds were in the middle of a group of thresholds with consistently high specificity and sensitivity. In the task of identifying simulated resilience SNPs, the selected model had a sensitivity of 0.95 and a specificity of 0.99. The hyperparameters selected for this model are shown in Supplementary Table 3. The model had an average AUC of 0.80 (95% CI: 0.77 – 0.83) across 5 trials in the task of classifying cases and controls in the high-risk subgroup and had an average AUC of 0.66 (95% CI: 0.56 – 0.76) in the same 5 trials in the task of classifying cases and controls in the low-risk subgroup, indicating that the model was successful in finding patterns that were more predictive or exclusively in the high-risk subgroup.

### Alzheimer’s Disease Analyses

We created risk-matched subgroups using the thresholds determined by the simulated data analyses. A logistic regression using PRS-CS had an AUC of 0.69 (95% CI: 0.67 – 0.71) in the independent validation subset. After matching based on PRS-CS, the same logistic regression model was no longer significantly predictive, with an AUC of 0.51 (95% CI: 0.46 – 0.55). The resulting high-risk subgroup contained 225 cases and 225 controls in the training subset and 106 cases and 106 controls in the validation subset. The low-risk subgroup contained 538 cases and 538 controls in the training subset and 227 cases and 227 controls in the validation subset. Age, sex, and *APOE-*ε*4* haplotype presence were not significantly different between low-risk and high-risk subgroups in cases or controls. Information about the age, sex, and *APOE-*ε*4* haplotype presence for cases and controls in each subgroup is available in Supplementary Table 4. After clumping, there were 919 SNPs (831 from risk GWAS and 88 from resilience GWAS) remaining as input into the neural network.

The best set of hyperparameters for the model trained on the training subset of these data is shown in Supplementary Table 3. Across ten trials, the model had an average AUC of 0.69 (95% CI: 0.68 – 0.69) in the task of classifying cases and controls in the high-risk subgroup and an average AUC of 0.53 (95% CI 0.52 – 0.55) in the task of classifying cases and controls in the low-risk subgroup. Feature-importance analysis found significant attribution differences in the 5 SNPs shown in Table 1 after Bonferroni correction for multiple-testing in high-risk cases and controls. Feature-importance analysis results for all SNPs used by our models alongside GWAS results for those SNPs are shown in Supplementary Table 5.

**Table 1:** Significant results from feature importance analysis. P-values were corrected using Bonferroni multiple-testing correction. Clumped SNPs is the number of correlated SNPs clumped into each primary SNP. Location represents the chromosome and position of the SNP.

| SNP | P-value | Clumped SNPs | Location |
| --- | --- | --- | --- |
| rs429358 | $1.9 \times 10^{-7}$ | 27 | 19:45411941 |
| rs12721051 | $5.9 \times 10^{-7}$ | 2 | 19:45422160 |
| rs12972156 | $6.3 \times 10^{-5}$ | 21 | 19:45387459 |
| rs10119 | 0.002 | 3 | 19:45406673 |
| rs2394936 | 0.037 | 8 | 7:98413656 |

The resilience score had a correlation of −0.09 (p = 1.4 × 10^-5^) with PRS-CS in the validation subset. After residualization, the two were not significantly correlated. In a logistic regression predicting case/control status, PRS-CS, the residualized resilience score, and their interaction were all significantly associated with case/control status. The results of this model are presented in Table 2. The interaction between PRS-CS and the residualized resilience score is visualized in Figure 3, demonstrating an increase in association between PRS-CS and case/control prediction as the resilience score decreases. The association between PRS-CS and case/control prediction was approximately twice as strong in the bottom 25% resilience score bin compared to the top 25% resilience score bin, with logistic regression coefficients of 8.3 × 10^5^ and 4.1 × 10^5^, respectively.

**Figure 3:**
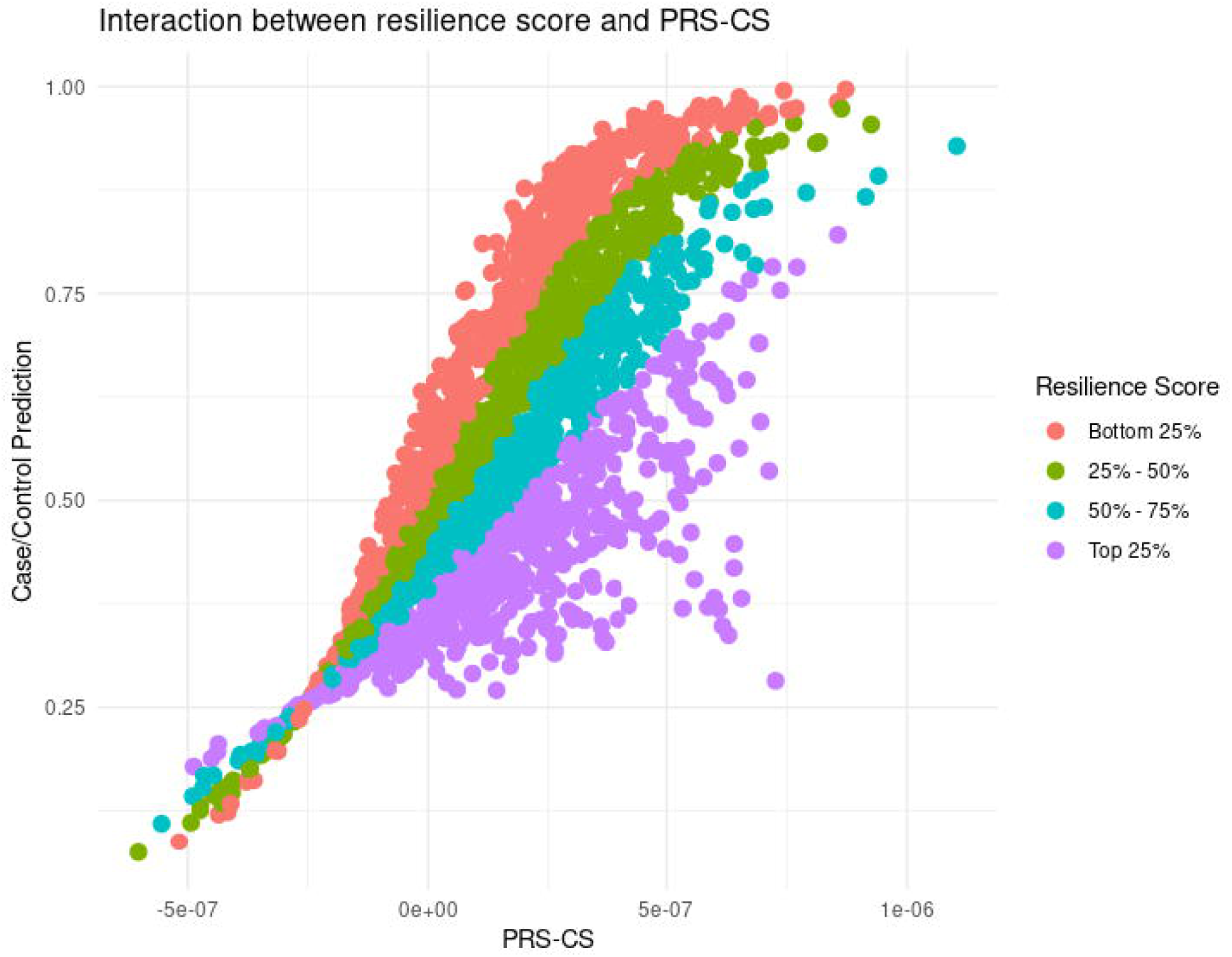
Interaction between resilience score and PRS-CS. The interaction between PRS-CS and the residualized resilience score visualized by binning residualized resilience score into 4 bins and plotting PRS-CS vs diagnosis prediction.

**Table 2.** Logistic regression predicting case/control status using PRS-CS, resilience score, and the interaction between PRS-CS and resilience score.

| Feature | Coefficient | Std. Error | Z | p |
| --- | --- | --- | --- | --- |
| PRS-CS | $1.1 \times 10^7$ | $8.4 \times 10^5$ | 13.1 | $< 2 \times 10^{-16}$ |
| Residualized resilience score | -1.9 | 0.2 | -8.8 | $< 2 \times 10^{-16}$ |
| PRS-CS * residualized resilience score | $-7.5 \times 10^6$ | $7.8 \times 10^5$ | -9.7 | $< 2 \times 10^{-16}$ |

\*: interaction

## Discussion

In both simulated data and actual Alzheimer’s disease data, our findings show that our adversarial multi-task neural network architecture improves our ability to detect markers of resilience, even when they are in linkage disequilibrium with risk alleles. We are the first to apply adversarial learning in the context of resilience to effectively unlearn patterns that are present in low-risk groups to explicitly drive the model towards learning the patterns that are specific to high-risk, resilient controls relative to risk-matched cases. This feature of the model is critical to ensuring that resilience effects are distinct from risk. Rather, the impact of resilience is to modify the effect of risk variants. In simulated data, these methods resulted in more consistent and specific determination of resilience SNPs compared to a neural network using a more traditional approach to capturing resilience effects. In AD data, we trained a model that was significantly more accurate at classifying cases and controls in the high-risk subset compared to the low-risk subset. We found 5 SNPs to be significantly implicated in this high-risk-specific model, suggesting these SNPs are markers of resilience in these high-risk controls, and not simply unaccounted for risk variants.

In a dataset of 1 million simulated individuals, we aimed to test how well our neural network architecture could separate the effects of resilience SNPs from the effects of other types of SNPs. As shown in Figure 2, we found that, compared with the standard approach of only using the high-risk subset to train the model, the specificity of our adversarial method in determining resilience SNPs was much higher in most of the sets of thresholds used to determine high-risk and low-risk subsets.

Perhaps most importantly, considering the noisier risk patterns in real data, the results of the adversarial models were much more consistent than the standard models across different conditions. This is critical because other data sets will inevitably differ in risk patterns and ideal thresholding criteria, so knowing that our method for detecting resilience markers performs consistently across different thresholds and model architectures is important towards trusting the output outside of our simulated dataset. The models for each threshold set were optimized in the same way and the results suggest that in the standard approach the sensitivity and specificity are dependent upon the best set of hyperparameters. In comparison, the high consistency of the adversarial approach suggests that the sensitivity and specificity are robust to different choices of model hyperparameters. In real-world data we rarely know the ground truth for which SNPs are markers of resilience and which SNPs represent additive risk, which makes it impossible to objectively select thresholds in the same way we were able to with simulated data. Our simulation results demonstrate that, across a range of parameter choices, the model is unlikely to produce a substantial number of false positives, providing reassurance that any significant SNPs can be interpreted as resilience-related variants.

The significant associations of PRS-CS and resilience score in the logistic regression model predicting AD diagnosis suggest that the effects of resilience score are in part independent of the effects of polygenic risk-scoring. The significant interaction between resilience score and PRS-CS, shown in Table 2, suggests that the resilience score is protective against polygenic risk by attenuating its effect. As shown in Figure 3, the effect of PRS-CS on AD diagnosis prediction was lower in groups with higher resilience scores. Conversely, resilience effects were more apparent at higher levels of risk. This was an expected result, since we anticipated that unaffected individuals with higher risk likely had resilience factors in place that counteracted their elevated risk to prevent development of the disorder. This result also aligns with a GWAS-oriented resilience-scoring method applied to AD that found higher resilience scores were associated with a lower risk of AD.(17) Our models were designed to detect resilience features and are not designed to maximize AUC. Since our adversarial learning approach is purposefully disruptive to the normal machine learning process, our models’ AUCs are expectedly lower than previous work that focused on maximizing classification accuracy.(29, 30) Rather, we impose a constraint of maintaining high specificity such that we are not simply (re)identifying risk SNPs. However, the difference between the AUC in the high-risk subgroup (0.69) and the AUC in the low-risk subgroup (0.53) across ten trials further suggests our model can find patterns that are more predictive of case/control status in the high-risk subgroup. This AUC difference mirrors the same difference seen in our models on simulated data that detected resilience SNPs with high specificity.

Our approach identified 5 SNPs that had significantly different feature-attributions between cases and controls. The SNPs were rs429358, rs12721051, rs12972156, rs10119, and rs2394936. The SNPs themselves are part of SNP clumps that we formed based on LD, so the results could represent those SNPs acting as markers for nearby causal resilience SNPs as well. The most statistically significant difference was seen in rs429358, one of the two SNPs that defines *APOE* variants. The *APOE* interaction pair, while known, is exactly the type of effect we would expect this analysis to identify. So, while our identification of rs429358 is not novel, its detection validates our methodology. This, by analogy, lends credibility to the other resilience SNPs detected via the same method. One of these is rs12721051 in the Apolipoprotein C1 (*APOC1*) gene. Multiple studies(11, 12) (7, 13)have found associations between *APOC1* and AD and interactions between in *APOC1* and *APOE*. Cudaback *et al*.(7) showed that *APOC1* is an *APOE*-genotype-dependent suppressor of glial activation. Zhou *et al*.(13) found that an *APOC1* insertion allele increased AD risk in *APOE*-ε4 carriers but did not increase risk in non-carriers.

Another resilience-associated SNP was rs12972156 located in the Nectin Cell Adhesion Molecule 2 (*NECTIN2*) gene. A study using latent class analysis found that *NECTIN2* was differentially associated with cognitive decline in three latent classes,(31) which offers evidence of conditional risk effects we would expect to see with risk/resilience interactions. rs10119 is in the translocase of outer mitochondrial membrane 40 (*TOMM40*) gene. *TOMM40* has been found to influence AD risk both independently(14) and in combination(14) with *APOE*. Zhu *et al*.(15) found that *TOMM40* and *APOE* variants synergistically increase the risk for AD.(15) One study found that the four genes implicated by our work, *NECTIN2*, *APOE*, *APOC1*, and *TOMM40*, were likely the genes affecting plasma *APOE* expression levels.(16) While the other SNPs identified by our method are well studied and within *APOE* and it’s flanking region on chromosome 19, rs2394936 is located on chromosome 7 and little is known about the SNP. The abundance of existing evidence supporting interactions between the SNPs and genes identified by our method adds validity to the output of our models and strengthens our interest in further studying and validating rs2394936.

While one of the two SNPs that defines *APOE* variants was among the identified SNPs, the other, rs7412, was not. We suspect that this is a result of the adversarial learning process purposefully driving down the weights of rs7412 to avoid using the *APOE*-ε4 haplotype as a feature. As shown in Supplementary Table 4, the proportion of *APOE*-ε4 carriers in controls is not different between the low-risk and high-risk subgroups (27.3% vs 27.4%). The proportion of *APOE*-ε4 carriers in cases between the low-risk and high-risk subgroups are slightly, but not significantly, different (60.4% vs 68.9%). Since *APOE*-ε4 appears to be strongly and similarly predictive in the high-risk and low-risk subgroups, the model drives weights away from that interaction. This reflects a limitation of our models; by purposefully avoiding features that are predictive in the low-risk subgroup to avoid false positives and detect features that are independent of log-additive risk, the model is unable to detect any real resilience features that are similarly predictive in the high-risk and low-risk subgroups.

Other limitations should also be considered. The resilience against clinical dementia seen in high-risk controls is likely to reflect some combination of biological and environmental factors. In our study, we address only the genomic part of potential biological resilience factors. Collecting and modeling multi-omic data may detect more forms of resilience and interactions between risk and resilience. We also did not have data on AD-related pathological burden or on the mechanism underlying resilience. Even if two individuals have the same resilience-associated SNPs, they might still differ in being classified as case or control if one had much greater AD-related pathology at the time of assessment. Resilience-associated SNPs may confer resilience against AD pathology that has already developed, or they may confer resilience against the effects of other genes that in turn reduce or prevent development of AD pathology in the first place. Our study was likely limited by the size of our AD data set. This is especially true in the *APOE*-ε4 carrier sub-analysis, which failed to produce a reproducible model with our limited data size. Likewise, the number of SNPs we used in our models, which we minimized to balance sample size and model complexity, likely limited our results by excluding SNPs that would have been identified as important. It’s possible that with larger data sets and more input SNPs, more markers of resilience may be identified.

In summary, we have described a novel multi-task, adversarial neural network method for identifying and combining markers of genomic resilience. The results of applying our method to a large, simulated data set suggest that the method is an improvement over prior work that will minimize false-positives. Applied to Alzheimer’s disease data, our approach produced resilience scores that were protective against polygenic risk and identified 5 SNPs as markers of resilience. This approach shows promise in identifying variants that mark areas of importance in understanding the pathology of AD, with the ultimate goal of improved diagnosis and prevention.

## Supporting information

ML Resilience Supplement

## Data Availability

Each simulated individual standard additive risk and total risk scores, and information on each simulated SNP are available in the supplementary data file

## Acknowledgements

Eric J. Barnett, Jiahui Hou, and Peter Holmans have no acknowledgments to disclose. Dr. Jonathan Hess’s research over the past two years has been supported by NIH/NINDS grant R01NS128535 and the CNY Community Foundation. Dr. Valentina Escott-Price’s research over the past two years has been supported by the UK Dementia Research Institute [UK DRI-3206] through UK DRI Ltd, principally funded by the Medical Research Council. Dr. Christine Fennema-Notestine’s research over the past two years has been supported by NIH grants R01AG064955, R01AG076838, R24MH129166, R01MH098742, R01 MH128887, P30MH062512, R01MH125720, R01 DA047906, R01DA047879, and P01 AG055367; and NIH contract 75N95023C00013. Dr. William Kremen’s research over the past two years has been supported by NIH/NIA grant R01AG050595, R01AG076838, R01AG064955, R01AG060470, and P01AG055367. Dr. Shu-Ju Lin’s research over the past two years is supported by NIH/NIA grants R01AG064955, R01AG050595, and R01AG076838. Dr. Chunling Zhang’s research over the past two years has been supported by NIH grants: R01AG064955, R01MH126459, R01NS128535. Dr. Chris Gaiteri’s research over the past two years has been supported by NIA and NIH grants R01AG061800, R01AG061798, U01AG079847. Dr. Jeremy Elman’s research over the past two years has been supported by NIH/NIA grants K01AG063805, R01AG076838 and R01AG050595. Dr. Stephen Faraone’s research over the past two years has been supported by the European Union’s Horizon 2020 research and innovation programme under grant agreement 965381; NIH/NIMH grants U01AR076092, R01MH116037, 1R01NS128535, R01MH131685, 1R01MH130899, U01MH135970, Massachusetts General, Otsuka, Corium Pharmaceuticals, Tris Pharmaceuticals, Oregon Health & Science University, Supernus Pharmaceuticals. His continuing medical education programs are supported by Corium Pharmaceuticals, Noven, Supernus Pharmaceuticals, Tris Pharmaceuticals, and The Upstate Foundation. **Dr. Stephen Glatt’s** research over the past two years has been supported by grant R01AG064955 from the U.S. National Institute on Aging and grant R21MH126494 from the U.S. National Institute of Mental Health.

