## Supplementary material for "A Novel Method to Disentangle Tightly Linked Risk and Resilience Genes for Brain Disorders: Application to Alzheimer’s Disease": ML Resilience Supplement

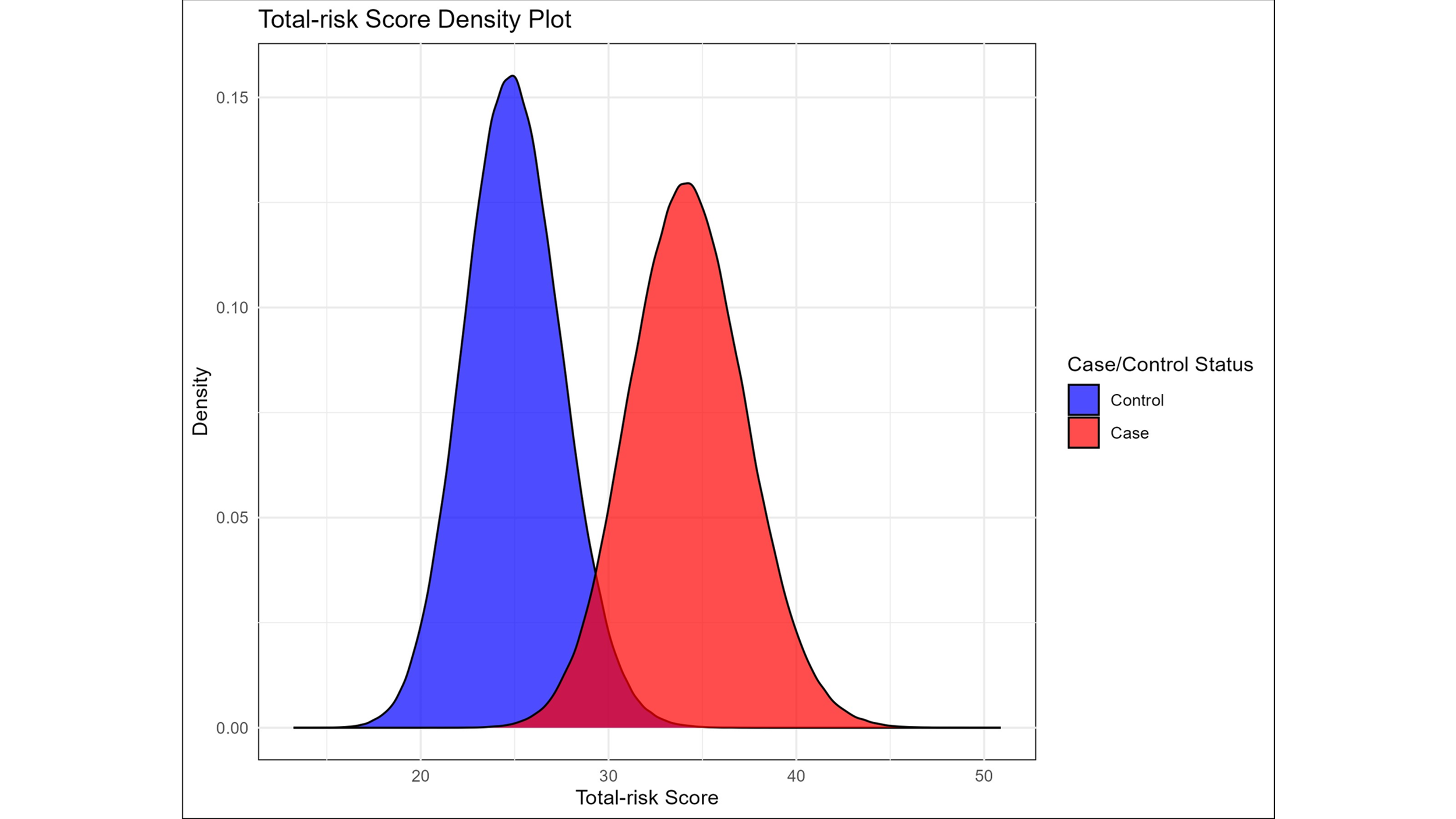

Supplementary Figure 1: Total-risk Score Density Plot. A density plot showing the distribution of cases and controls based on total-risk score.

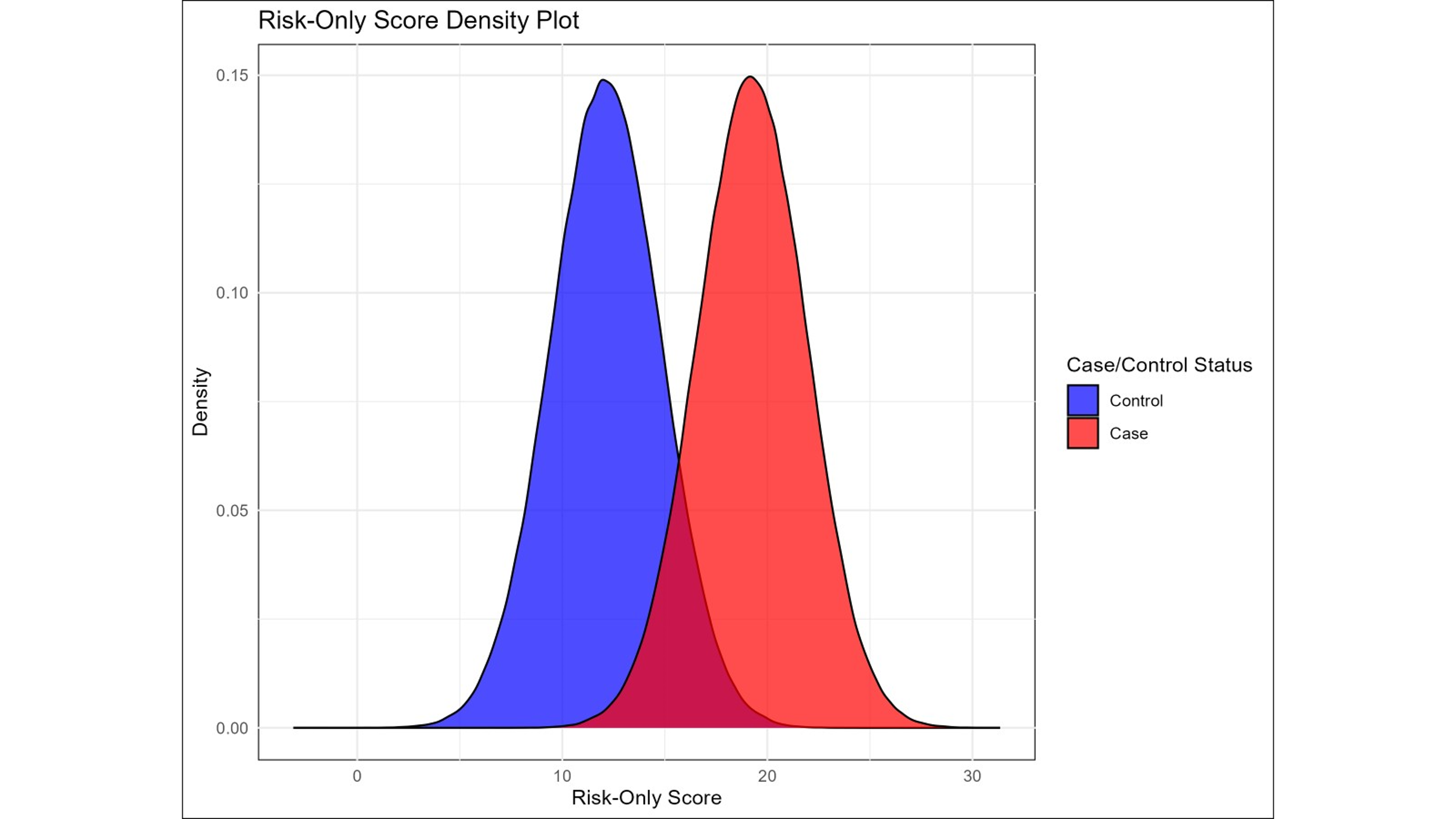

Supplementary Figure 2: Risk-Only Score Density Plot. A density plot showing the distribution of cases and controls based on risk-only score.

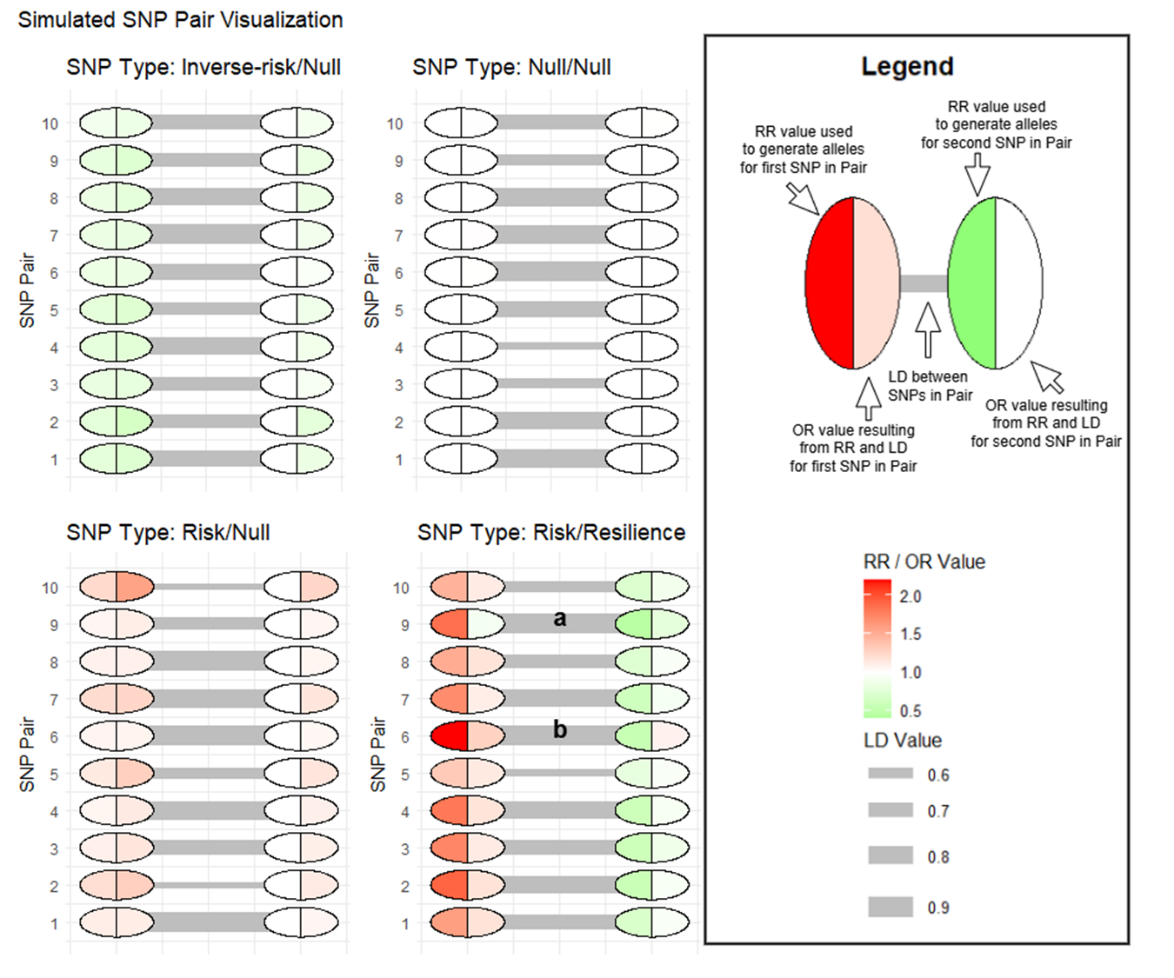

**Supplementary Figure 3: Simulated SNP-Pair Visualization**. Visualization of the first ten SNP-pairs for each SNP-pair type. For each SNP-pair, the left circle represents the first SNP named in the pair type name (for example the risk SNP in the risk/null pair). The right circle represents the second SNP named in the pair type name (for example the null SNP in the risk/null pair). The left side of each circle represents the relative risk (RR) value used to generate alleles for that SNP, while the right side of each circle represents the odds ratio (OR) that resulted for that SNP based on RR, allele frequency, and LD of both SNPs in the pair. This illustrates the tendency of simulated Null SNPs to look like the type of SNP with which they are in LD. It also illustrates the tendency of risk/resilience pairs to reduce the apparent effect size of correlated SNPs with opposing effects. Marker (a) shows a risk/resilience SNP-pair where the strong LD and effect size of the resilience SNP nullifies the effect of the risk SNP, resulting in the risk SNP having an OR very close to 1. This risk SNP would likely be missed in GWAS, but in those without the closely linked resilience SNP would be an important risk factor. Marker (b) shows a risk/resilience SNP-pair where the strong LD and effect size of the risk SNP makes the resulting OR of the resilience SNP appear as though it is a risk SNP in GWAS, illustrating the importance of understanding these potential interactions.

| **Supplementary Table 1.** Hyperparameter Optimization Ranges | |
| --- | --- |
| Optimization Task | Optimization Range |
| Epochs | 10 - 1000 |
| Nodes Within Dense Layer | 10 - 100 |
| L1 regularization factor | 1e-5 – 1e-3 |
| Dropout Rate (AD only) | 0 – 0.7 |
| Gradient Reversal Weight | 0 – 3 |
| Learning Rate | 1e-5 – 1e-1 |

| **Supplementary Table 2:** Thresholding Analysis Results | | | | | | |
| --- | --- | --- | --- | --- | --- | --- |
| High risk lower threshold | Low risk lower threshold | Low risk upper threshold | Adversarial model sensitivity | Adversarial model specificity | Non adversarial model sensitivity | Non adversarial model specificity |
| 0.7 | 0.2 | 0.3 | 0.920245 | 0.981183 | 1 | 0.057796 |
| 0.7 | 0.2 | 0.4 | 0.969325 | 0.967294 | 0.969325 | 0.993728 |
| 0.7 | 0.2 | 0.5 | 0.981595 | 0.959677 | 0.779141 | 0.273746 |
| 0.7 | 0.2 | 0.6 | 0.969325 | 0.948477 | 1 | 0.005824 |
| 0.7 | 0.2 | 0.7 | 0.595092 | 0.985215 | 1 | 0.036738 |
| 0.7 | 0.4 | 0.5 | 0.97546 | 0.978047 | 0.582822 | 0.414427 |
| 0.7 | 0.4 | 0.6 | 0.981595 | 0.982527 | 0.220859 | 0.257168 |
| 0.7 | 0.4 | 0.7 | 0.97546 | 0.971326 | 0.993865 | 0.290771 |
| 0.7 | 0 | 0.3 | 0.932515 | 0.573029 | 0.993865 | 0.923835 |
| 0.7 | 0 | 0.4 | 0.957055 | 0.922939 | 0.993865 | 0.428763 |
| 0.7 | 0 | 0.5 | 0.92638 | 0.783602 | 0.92638 | 0.37052 |
| 0.7 | 0 | 0.6 | 0.981595 | 0.983871 | 1 | 0.002688 |
| 0.7 | 0 | 0.7 | 0.981595 | 0.978495 | 0.742331 | 0.282258 |
| 0.8 | 0.2 | 0.3 | 0.809816 | 0.623208 | 0.98773 | 0.005824 |
| 0.8 | 0.2 | 0.4 | 0.97546 | 0.966398 | 0.276074 | 0.375896 |
| 0.8 | 0.2 | 0.5 | 0.96319 | 0.973118 | 1 | 0.000448 |
| 0.8 | 0.2 | 0.6 | 0.96319 | 0.96371 | 1 | 0.060036 |
| 0.8 | 0.2 | 0.7 | 0.944785 | 0.74328 | 0.993865 | 0.338262 |
| 0.8 | 0.4 | 0.5 | 0.981595 | 0.981631 | 0.349693 | 0.342742 |
| 0.8 | 0.4 | 0.6 | 0.96319 | 0.970878 | 1 | 0 |
| 0.8 | 0.4 | 0.7 | 0.96319 | 0.967294 | 1 | 0.045251 |
| 0.8 | 0 | 0.3 | 0.96319 | 0.939516 | 1 | 0.601703 |
| 0.8 | 0 | 0.4 | 0.95092 | 0.957437 | 1 | 0.136201 |
| 0.8 | 0 | 0.5 | 0.969325 | 0.980735 | 1 | 0.33871 |
| 0.8 | 0 | 0.6 | 0.98773 | 0.985663 | 1 | 0.000448 |
| 0.8 | 0 | 0.7 | 0.981595 | 0.952957 | 0.834356 | 0.044803 |
| 0.9 | 0.2 | 0.3 | 0.95092 | 0.996864 | 0.993865 | 0.215502 |
| 0.9 | 0.2 | 0.4 | 0.944785 | 0.994624 | 0.993865 | 0.354391 |
| 0.9 | 0.2 | 0.5 | 0.95092 | 0.992384 | 1 | 0.327509 |
| 0.9 | 0.2 | 0.6 | 0.957055 | 0.991039 | 1 | 0.054211 |
| 0.9 | 0.2 | 0.7 | 0.932515 | 0.990591 | 0.97546 | 0.041667 |
| 0.9 | 0.4 | 0.5 | 0.944785 | 0.992384 | 0.03681 | 0.285394 |
| 0.9 | 0.4 | 0.6 | 0.95092 | 0.976254 | 0.472393 | 0.202061 |
| 0.9 | 0.4 | 0.7 | 0.96319 | 0.992384 | 1 | 0.012545 |
| 0.9 | 0 | 0.3 | 0.889571 | 0.998656 | 0.705521 | 0.761201 |
| 0.9 | 0 | 0.4 | 0.957055 | 0.985215 | 1 | 0.390233 |
| 0.9 | 0 | 0.5 | 0.944785 | 0.999104 | 0.98773 | 0.008961 |
| 0.9 | 0 | 0.6 | 0.93865 | 0.797491 | 1 | 0 |
| 0.9 | 0 | 0.7 | 0.95092 | 0.988799 | 1 | 0.118728 |
| 0.95 | 0.2 | 0.3 | 0.981595 | 0.903674 | 0.159509 | 0.469534 |
| 0.95 | 0.2 | 0.4 | 0.840491 | 0.988351 | 0.98773 | 0.5569 |
| 0.95 | 0.2 | 0.5 | 0.766871 | 0.875448 | 0.042945 | 0.355735 |
| 0.95 | 0.2 | 0.6 | 0.828221 | 0.999552 | 0.245399 | 0.480287 |
| 0.95 | 0.2 | 0.7 | 0.822086 | 0.999104 | 0.98773 | 0.27733 |
| 0.95 | 0.4 | 0.5 | 0.803681 | 0.999552 | 0.484663 | 0.663082 |
| 0.95 | 0.4 | 0.6 | 0.834356 | 0.919355 | 0.171779 | 0.263889 |
| 0.95 | 0.4 | 0.7 | 0.858896 | 0.994176 | 0.981595 | 0.34991 |
| 0.95 | 0 | 0.3 | 0.711656 | 1 | 0.791411 | 0.934588 |
| 0.95 | 0 | 0.4 | 0.748466 | 0.920699 | 1 | 0.204301 |
| 0.95 | 0 | 0.5 | 0.766871 | 0.992832 | 0.754601 | 0.585573 |
| 0.95 | 0 | 0.6 | 0.791411 | 0.999104 | 1 | 0.141577 |
| 0.95 | 0 | 0.7 | 0.754601 | 0.993728 | 0.883436 | 0.309588 |

| **Supplementary Table 3.** Model Hyperparameters | |  |
| --- | --- | --- |
| Hyperparameter | Simulation Model | AD Model |
| Layers | 1 | 1 |
| Epochs | 313 | 776 |
| Nodes Within Dense Layer | 27 | 98 |
| L1 regularization factor | 6.4e-6 | 0.0009 |
| Dropout Rate | --- | 0.39 |
| Gradient Reversal Weight | 0.35 | 0.22 |
| Learning Rate | 0.0094 | 0.0097 |

| **Supplementary Table 4.** Age, Sex, and APOE-ε4 Haplotype Presence in Validation Subset Cases and Controls | | | |
| --- | --- | --- | --- |
|  | Low-risk subgroup | High-risk subgroup | p |
| Mean age: controls | 75.9 years | 76.0 years | 0.94 |
| Mean age: cases | 73.3 years | 72.7 years | 0.48 |
| Percent female: controls | 65.6% | 65.1% | 1 |
| Percent female: cases | 48.9% | 49.1% | 1 |
| Percent APOE-ε4 carriers: controls | 27.3% | 27.4% | 1 |
| Percent APOE-ε4 carriers: cases | 60.4% | 68.9% | 0.17 |

| **Supplementary Table 5**: Feature Importance and GWAS Results for all modeled SNPs | | | | | |
| --- | --- | --- | --- | --- | --- |
| SNP | Bonferroni_adjusted_ML_feature_importance_p_value | CHR | BP | A1 | GWAS P |
| rs429358 | 1.91E-07 | 19 | 45411941 | C | 2.66E-09 |
| rs12721051 | 5.91E-07 | 19 | 45422160 | G | 8.97E-07 |
| 19:45387459:C:G | 6.26E-05 | 19 | 45387459 | G | 4.33E-05 |
| rs10119 | 0.002002469 | 19 | 45406673 | A | 2.50E-05 |
| rs2394936 | 0.037607948 | 7 | 98413656 | C | 5.77E-05 |
| rs439401 | 0.09915937 | 19 | 45414451 | T | 0.000284 |
| rs12974310 | 0.158261097 | 19 | 46161804 | G | 0.5208 |
| rs74236526 | 0.158261097 | 12 | 72015382 | A | 0.8001 |
| 9:21432043:A:T | 0.16300939 | 9 | 21432043 | T | 0.7368 |
| rs1160985 | 0.16300939 | 19 | 45403412 | T | 0.03406 |
| rs71352239 | 0.19832089 | 19 | 45429543 | T | 0.5055 |
| rs147711004 | 0.208521767 | 19 | 45337918 | A | 0.8327 |
| rs3819121 | 0.208521767 | 2 | 1.36E+08 | T | 0.1046 |
| rs3925681 | 0.23294932 | 19 | 45421100 | A | 0.01719 |
| rs77911607 | 0.261986781 | 8 | 1.45E+08 | T | 0.7426 |
| rs157580 | 0.264070631 | 19 | 45395266 | G | 0.001075 |
| 19:45222789:G:C | 0.294297523 | 19 | 45222789 | C | 0.59 |
| rs10845383 | 0.294297523 | 12 | 11747950 | T | 0.9089 |
| rs11882134 | 0.294297523 | 19 | 45123303 | A | 0.3305 |
| rs2965156 | 0.294297523 | 19 | 45188429 | C | 0.4607 |
| rs12983547 | 0.342539138 | 19 | 45091007 | A | 0.8587 |
| rs4147910 | 0.342539138 | 19 | 1047078 | G | 0.3694 |
| 6:31134888:T:C | 0.41551713 | 6 | 31134888 | T | 0.9618 |
| rs10077003 | 0.41551713 | 5 | 1.3E+08 | T | 0.7388 |
| rs12906722 | 0.41551713 | 15 | 58692715 | T | 0.52 |
| rs13207334 | 0.41551713 | 6 | 47383890 | G | 0.5395 |
| rs150943557 | 0.41551713 | 2 | 2.34E+08 | G | 0.2109 |
| rs1968449 | 0.41551713 | 19 | 45544351 | T | 0.3153 |
| rs2071293 | 0.41551713 | 6 | 32062687 | A | 0.715 |
| rs4602803 | 0.41551713 | 7 | 1.24E+08 | G | 1.13E-05 |
| rs7721972 | 0.431552171 | 5 | 1.3E+08 | G | 0.1845 |
| rs4335021 | 0.44241356 | 6 | 32386619 | T | 0.4212 |
| 13:19772942:C:T | 0.459731188 | 13 | 19772942 | C | 9.83E-05 |
| rs10808204 | 0.459731188 | 7 | 1.24E+08 | T | 1.51E-06 |
| rs12590654 | 0.459731188 | 14 | 92938855 | A | 0.9431 |
| rs585487 | 0.459731188 | 19 | 367313 | G | 0.01098 |
| rs7731850 | 0.459731188 | 5 | 1.3E+08 | G | 0.3768 |
| rs9592520 | 0.459731188 | 13 | 68181162 | G | 1.75E-05 |
| rs2394447 | 0.460904567 | 6 | 30826835 | T | 0.8119 |
| rs35853021 | 0.460904567 | 15 | 58680643 | T | 0.1056 |
| rs889555 | 0.460904567 | 16 | 31122571 | T | 0.2218 |
| rs406456 | 0.471886787 | 19 | 45382717 | G | 0.1857 |
| rs4727449 | 0.471886787 | 7 | 99785750 | T | 0.152 |
| rs9304690 | 0.485164395 | 19 | 50453317 | T | 0.09166 |
| rs12150984 | 0.499457001 | 19 | 45202027 | A | 0.5288 |
| rs2307418 | 0.499457001 | 1 | 1.61E+08 | G | 0.7851 |
| rs36051450 | 0.499457001 | 11 | 1.23E+08 | C | 0.7655 |
| rs55808779 | 0.499457001 | 11 | 85873085 | T | 0.9223 |
| rs6949267 | 0.499457001 | 7 | 1.43E+08 | C | 0.1958 |
| rs6953295 | 0.499457001 | 7 | 1.23E+08 | T | 2.68E-05 |
| rs7176404 | 0.499457001 | 15 | 64376238 | C | 0.03427 |
| rs7221751 | 0.499457001 | 17 | 1393143 | A | 0.5579 |
| rs420332 | 0.514268121 | 16 | 89698752 | T | 0.3552 |
| 19:45534041:C:T | 0.544395996 | 19 | 45534041 | T | 0.2011 |
| rs1390425 | 0.544395996 | 15 | 34658419 | G | 4.89E-05 |
| rs3129731 | 0.544395996 | 6 | 32682207 | T | 0.6675 |
| rs4451810 | 0.544395996 | 12 | 26404284 | G | 0.535 |
| rs73050205 | 0.544395996 | 19 | 45356464 | A | 0.7438 |
| rs871269 | 0.544395996 | 5 | 1.5E+08 | T | 0.8097 |
| rs9275214 | 0.546373021 | 6 | 32658665 | A | 0.3437 |
| 19:45200681:G:T | 0.567947626 | 19 | 45200681 | T | 0.4944 |
| rs62052663 | 0.567947626 | 16 | 90153346 | A | 0.7427 |
| 12:123309475:T:C | 0.574756963 | 12 | 1.23E+08 | T | 0.9549 |
| rs73156698 | 0.574756963 | 7 | 99573259 | G | 0.7897 |
| rs115004308 | 0.576845892 | 1 | 2.26E+08 | A | 0.5166 |
| rs6678085 | 0.577020936 | 1 | 22508799 | T | 1.75E-05 |
| rs112972879 | 0.60432628 | 19 | 46165082 | A | 0.02451 |
| 16:30878366:C:T | 0.630859962 | 16 | 30878366 | C | 0.07997 |
| rs7160605 | 0.630859962 | 14 | 92993336 | C | 0.3751 |
| rs2965163 | 0.637054695 | 19 | 45200526 | A | 0.2533 |
| rs9271366 | 0.637653335 | 6 | 32586854 | G | 0.4521 |
| 7:15302128:C:T | 0.63868625 | 7 | 15302128 | C | 7.10E-05 |
| rs1135062 | 0.63868625 | 19 | 45322744 | G | 0.2782 |
| rs7182844 | 0.63868625 | 15 | 74673971 | T | 0.1141 |
| rs754340 | 0.63868625 | 19 | 45163671 | C | 0.864 |
| rs78190442 | 0.63868625 | 8 | 17358859 | C | 1.65E-05 |
| rs3752246 | 0.642682629 | 19 | 1056492 | G | 0.4244 |
| rs3873453 | 0.642682629 | 6 | 32683382 | T | 0.9144 |
| rs7254776 | 0.642682629 | 19 | 45227742 | C | 0.02754 |
| rs753127 | 0.642682629 | 14 | 92857300 | C | 0.6573 |
| rs920916 | 0.642682629 | 15 | 58688477 | A | 0.002762 |
| 17:63816298:C:T | 0.650783461 | 17 | 63816298 | T | 8.45E-05 |
| rs11669338 | 0.650783461 | 19 | 45382984 | G | 0.5216 |
| rs35961830 | 0.650783461 | 16 | 31032317 | C | 0.7625 |
| rs585046 | 0.650783461 | 20 | 1690506 | T | 0.2726 |
| rs6122775 | 0.650783461 | 20 | 36239111 | A | 0.7905 |
| rs62119263 | 0.650783461 | 19 | 45125197 | C | 0.7349 |
| rs79228497 | 0.650783461 | 5 | 73970106 | A | 0.9248 |
| rs12983944 | 0.657302626 | 19 | 45820238 | G | 0.9237 |
| rs78749443 | 0.65832163 | 19 | 1033190 | A | 0.5209 |
| 6:31439740:G:T | 0.658655433 | 6 | 31439740 | G | 0.06342 |
| rs10498633 | 0.658655433 | 14 | 92926952 | T | 0.2482 |
| rs11257190 | 0.658655433 | 10 | 11644108 | A | 0.6096 |
| rs11666329 | 0.658655433 | 19 | 45354296 | G | 0.4088 |
| rs11784619 | 0.658655433 | 8 | 1.45E+08 | A | 0.819 |
| rs16884295 | 0.658655433 | 5 | 10223028 | C | 0.4876 |
| rs1892676 | 0.658655433 | 21 | 27593540 | C | 0.0196 |
| rs2298813 | 0.658655433 | 11 | 1.21E+08 | A | 0.03819 |
| rs2965164 | 0.658655433 | 19 | 45202052 | T | 0.229 |
| rs35146386 | 0.658655433 | 7 | 1E+08 | A | 0.459 |
| rs62545043 | 0.658655433 | 9 | 74018099 | C | 0.7138 |
| rs72747098 | 0.658655433 | 15 | 63525249 | G | 0.04866 |
| rs9889994 | 0.675201868 | 17 | 47301268 | A | 0.08086 |
| rs4790160 | 0.677167577 | 17 | 1425357 | T | 0.8311 |
| 19:54814234:C:T | 0.690269221 | 19 | 54814234 | C | 0.6089 |
| rs911163 | 0.690269221 | 20 | 54985113 | A | 0.3204 |
| rs2079855 | 0.709478496 | 7 | 8107743 | A | 0.2744 |
| rs2596465 | 0.709478496 | 6 | 31412948 | T | 0.7432 |
| rs347117 | 0.709478496 | 15 | 59000957 | T | 0.5561 |
| rs55840414 | 0.709478496 | 19 | 45343579 | A | 0.3461 |
| rs6024397 | 0.709478496 | 20 | 54344199 | A | 5.41E-05 |
| rs6690215 | 0.709478496 | 1 | 2.08E+08 | C | 0.2407 |
| rs821226 | 0.709478496 | 16 | 79603512 | T | 0.8977 |
| rs10806425 | 0.718212083 | 6 | 90926612 | A | 0.0243 |
| rs1136207 | 0.718212083 | 1 | 1.61E+08 | T | 0.6913 |
| rs11637751 | 0.718212083 | 15 | 63643524 | A | 0.8267 |
| rs2361282 | 0.718212083 | 3 | 36722232 | A | 5.14E-05 |
| rs3129712 | 0.718212083 | 6 | 32657200 | A | 0.8898 |
| rs4835494 | 0.718212083 | 4 | 1.49E+08 | C | 0.325 |
| rs58124010 | 0.718212083 | 17 | 5139808 | T | 0.4265 |
| rs61821016 | 0.718212083 | 1 | 2.07E+08 | C | 0.7422 |
| rs6935940 | 0.718212083 | 6 | 32683691 | C | 0.4488 |
| rs75836995 | 0.718212083 | 2 | 1.28E+08 | C | 0.3152 |
| rs7699824 | 0.718212083 | 4 | 1.6E+08 | T | 0.6299 |
| rs6867080 | 0.722194039 | 5 | 1.3E+08 | C | 0.3253 |
| rs765982 | 0.722194039 | 7 | 14023807 | C | 0.4642 |
| rs9552391 | 0.736221348 | 13 | 21778534 | T | 0.2901 |
| 6:47432637:C:T | 0.741382975 | 6 | 47432637 | C | 0.3854 |
| rs10510066 | 0.741382975 | 10 | 1.22E+08 | T | 0.5832 |
| rs11576522 | 0.741382975 | 1 | 2.08E+08 | A | 0.9576 |
| rs11623883 | 0.741382975 | 14 | 92959216 | G | 0.5653 |
| rs11956806 | 0.741382975 | 5 | 1.72E+08 | A | 3.13E-05 |
| rs1532278 | 0.741382975 | 8 | 27466315 | T | 0.991 |
| rs17071872 | 0.741382975 | 13 | 80376988 | A | 0.7932 |
| rs2741342 | 0.741382975 | 8 | 27330096 | T | 0.04564 |
| rs28399635 | 0.741382975 | 19 | 45323243 | G | 0.5959 |
| rs2889414 | 0.741382975 | 19 | 45297928 | G | 0.2977 |
| rs58986836 | 0.741382975 | 8 | 1.44E+08 | C | 0.199 |
| rs6709535 | 0.741382975 | 2 | 1.26E+08 | A | 2.30E-05 |
| rs74960871 | 0.741382975 | 6 | 45706189 | A | 0.611 |
| rs9394764 | 0.741382975 | 6 | 41140984 | C | 0.6882 |
| rs9920849 | 0.741382975 | 15 | 92231370 | C | 0.05224 |
| 10:11723537:A:T | 0.747701588 | 10 | 11723537 | A | 0.2864 |
| 19:45583584:A:T | 0.747701588 | 19 | 45583584 | T | 0.8947 |
| 19:46052898:C:T | 0.747701588 | 19 | 46052898 | T | 0.7041 |
| 2:234070587:A:T | 0.747701588 | 2 | 2.34E+08 | T | 0.0587 |
| rs11234501 | 0.747701588 | 11 | 85696170 | A | 0.3302 |
| rs11668327 | 0.747701588 | 19 | 45398633 | C | 0.07147 |
| rs11879589 | 0.747701588 | 19 | 45373276 | A | 0.2307 |
| rs12212415 | 0.747701588 | 6 | 1.5E+08 | T | 0.8405 |
| rs1265762 | 0.747701588 | 6 | 32321115 | C | 0.06642 |
| rs16964189 | 0.747701588 | 15 | 51494237 | T | 4.74E-06 |
| rs201172 | 0.747701588 | 1 | 6719735 | T | 0.7986 |
| rs2074453 | 0.747701588 | 19 | 1080189 | C | 0.9782 |
| rs2632516 | 0.747701588 | 17 | 56409089 | C | 0.03862 |
| rs2638281 | 0.747701588 | 19 | 49210869 | A | 0.823 |
| rs2741341 | 0.747701588 | 8 | 27330286 | G | 0.8661 |
| rs283793 | 0.747701588 | 15 | 76774752 | G | 0.01583 |
| rs28399637 | 0.747701588 | 19 | 45324138 | A | 0.4006 |
| rs34883952 | 0.747701588 | 1 | 2.08E+08 | T | 0.3651 |
| rs35194062 | 0.747701588 | 19 | 45505177 | A | 0.7977 |
| rs365653 | 0.747701588 | 19 | 45361646 | G | 0.5816 |
| rs3758866 | 0.747701588 | 11 | 60047502 | T | 0.7045 |
| rs3857580 | 0.747701588 | 6 | 41216334 | G | 0.7563 |
| rs440277 | 0.747701588 | 19 | 45361224 | A | 0.879 |
| rs55802822 | 0.747701588 | 14 | 92952854 | G | 0.3898 |
| rs56124100 | 0.747701588 | 13 | 22429591 | T | 0.8038 |
| rs62117780 | 0.747701588 | 19 | 44752721 | G | 0.8027 |
| rs6934355 | 0.747701588 | 6 | 47362439 | C | 0.5064 |
| rs6979446 | 0.747701588 | 7 | 55015505 | T | 0.6157 |
| rs7096909 | 0.747701588 | 10 | 82284512 | A | 0.7546 |
| rs73357369 | 0.747701588 | 17 | 74685836 | A | 2.52E-06 |
| rs7659738 | 0.747701588 | 4 | 40303809 | T | 0.5268 |
| rs79076865 | 0.747701588 | 11 | 3103777 | G | 0.4959 |
| rs8104483 | 0.747701588 | 19 | 45372354 | G | 0.05663 |
| rs9427715 | 0.747701588 | 1 | 2.02E+08 | G | 1.43E-05 |
| rs965839 | 0.747701588 | 10 | 11718139 | C | 0.7523 |
| rs9900530 | 0.747701588 | 17 | 65493202 | A | 0.8202 |
| rs56176579 | 0.749229499 | 17 | 5262253 | T | 0.5216 |
| 1:207666061:C:T | 0.753928167 | 1 | 2.08E+08 | C | 0.4143 |
| rs11668972 | 0.753928167 | 19 | 45131677 | A | 0.5964 |
| rs661271 | 0.753928167 | 11 | 85653256 | T | 0.4658 |
| rs7926354 | 0.753928167 | 11 | 59962189 | A | 0.02638 |
| rs793571 | 0.753928167 | 15 | 59141706 | G | 0.3221 |
| 19:45264110:G:T | 0.754057234 | 19 | 45264110 | T | 0.3514 |
| rs1108047 | 0.754057234 | 22 | 40402817 | G | 5.11E-05 |
| rs17711542 | 0.754057234 | 19 | 43837552 | C | 0.9363 |
| rs2927468 | 0.754057234 | 19 | 45357939 | A | 0.8519 |
| rs4337014 | 0.754057234 | 11 | 7490797 | A | 8.85E-06 |
| rs714948 | 0.754057234 | 19 | 45165912 | A | 0.4051 |
| rs72835502 | 0.754057234 | 17 | 47453635 | A | 0.494 |
| rs11100203 | 0.755255928 | 4 | 1.6E+08 | G | 0.6116 |
| rs11669442 | 0.756322933 | 19 | 46374557 | C | 0.4984 |
| rs17128291 | 0.756322933 | 14 | 92882826 | G | 0.9627 |
| rs1782652 | 0.756322933 | 10 | 81074125 | A | 0.08186 |
| rs3792789 | 0.756322933 | 5 | 1.5E+08 | G | 0.9127 |
| rs7920721 | 0.756322933 | 10 | 11720308 | G | 0.9964 |
| rs9982724 | 0.756322933 | 21 | 40461378 | A | 0.7826 |
| 16:81903724:T:G | 0.761282293 | 16 | 81903724 | T | 0.04866 |
| rs2385089 | 0.761282293 | 19 | 18550434 | A | 0.1923 |
| rs3213934 | 0.76445835 | 11 | 85627039 | G | 0.4874 |
| rs12972892 | 0.768770076 | 19 | 45982373 | G | 0.3875 |
| rs3096145 | 0.768770076 | 16 | 23613000 | G | 0.9188 |
| rs660745 | 0.777062967 | 19 | 49219459 | T | 0.8515 |
| 14:92934509:A:G | 0.790647403 | 14 | 92934509 | A | 0.8258 |
| rs1946990 | 0.790647403 | 22 | 38912771 | G | 0.2745 |
| rs2242601 | 0.790647403 | 7 | 1.43E+08 | G | 0.4231 |
| rs72828895 | 0.790647403 | 17 | 4976660 | G | 0.8164 |
| 19:1039444:C:T | 0.7908952 | 19 | 1039444 | C | 0.08097 |
| 7:1593963:A:G | 0.7908952 | 7 | 1593963 | A | 0.1644 |
| rs10407439 | 0.7908952 | 19 | 45341948 | A | 0.759 |
| rs11039154 | 0.7908952 | 11 | 47278502 | T | 0.3051 |
| rs12042437 | 0.7908952 | 1 | 1.61E+08 | T | 0.3191 |
| rs1784920 | 0.7908952 | 11 | 1.21E+08 | A | 0.8603 |
| rs2018400 | 0.7908952 | 15 | 63626540 | T | 0.9904 |
| rs444694 | 0.7908952 | 19 | 44309076 | G | 0.4641 |
| rs62118504 | 0.7908952 | 19 | 45734751 | G | 0.2057 |
| rs77594976 | 0.7908952 | 19 | 44407177 | A | 0.6065 |
| rs11569565 | 0.791528309 | 19 | 6678474 | A | 6.91E-05 |
| rs3815170 | 0.791528309 | 19 | 1013148 | A | 0.0793 |
| rs1056434 | 0.79411286 | 6 | 47445789 | A | 0.3399 |
| 8:51652266:C:G | 0.798050513 | 8 | 51652266 | G | 0.1342 |
| rs12570047 | 0.798050513 | 10 | 11501699 | A | 0.787 |
| rs4575098 | 0.798050513 | 1 | 1.61E+08 | A | 0.8604 |
| 6:32266310:T:C | 0.799975019 | 6 | 32266310 | T | 0.5299 |
| rs6728532 | 0.800172201 | 2 | 2.08E+08 | C | 0.05823 |
| rs6859 | 0.800172201 | 19 | 45382034 | G | 0.007985 |
| rs2240643 | 0.800631592 | 7 | 1.56E+08 | C | 0.3527 |
| rs7257002 | 0.800631592 | 19 | 5043527 | T | 0.9574 |
| rs9268835 | 0.80183728 | 6 | 32428115 | A | 0.8763 |
| rs12549671 | 0.809937759 | 8 | 27500084 | G | 0.7446 |
| rs33817 | 0.809937759 | 5 | 1.04E+08 | A | 7.25E-05 |
| rs3098176 | 0.811210531 | 15 | 50781963 | A | 0.8459 |
| rs7260482 | 0.811210531 | 19 | 45143942 | C | 0.7772 |
| rs9469112 | 0.811210531 | 6 | 32415153 | T | 0.9803 |
| 19:45105867:A:G | 0.812904036 | 19 | 45105867 | G | 0.3074 |
| rs4889620 | 0.812904036 | 16 | 31131174 | A | 0.5483 |
| rs6130736 | 0.812904036 | 20 | 43668059 | T | 0.8247 |
| rs73217076 | 0.812904036 | 4 | 11049800 | A | 0.3554 |
| rs10761418 | 0.815958062 | 10 | 61592966 | A | 0.4327 |
| 4:109908570:C:T | 0.816838699 | 4 | 1.1E+08 | C | 0.6862 |
| 14:92916942:G:T | 0.820284646 | 14 | 92916942 | T | 0.2457 |
| rs13416785 | 0.820635821 | 2 | 2.03E+08 | G | 0.5725 |
| rs4141143 | 0.835369034 | 17 | 11256032 | T | 1.94E-05 |
| rs12796308 | 0.841709758 | 11 | 23343707 | A | 0.1083 |
| 8:104029528:A:G | 0.841773375 | 8 | 1.04E+08 | G | 0.09675 |
| rs2040102 | 0.84768743 | 1 | 12735511 | C | 0.699 |
| 14:73976934:T:C | 0.84950072 | 14 | 73976934 | C | 0.04161 |
| rs2440070 | 0.850783266 | 10 | 11488309 | C | 0.7797 |
| rs72959149 | 0.850783266 | 18 | 56598031 | G | 0.2516 |
| rs384653 | 0.858848756 | 19 | 45370335 | A | 0.6539 |
| 15:77256130:T:G | 0.859929802 | 15 | 77256130 | G | 0.04305 |
| rs10905989 | 0.862684812 | 10 | 11719593 | T | 0.6945 |
| 19:54814414:G:T | 0.865825003 | 19 | 54814414 | T | 0.2285 |
| rs10421247 | 0.865825003 | 19 | 45657486 | C | 0.541 |
| rs10484759 | 0.865825003 | 6 | 1.27E+08 | T | 0.1603 |
| rs111380074 | 0.865825003 | 20 | 55104505 | A | 0.4424 |
| rs11979746 | 0.865825003 | 7 | 1.24E+08 | A | 1.42E-05 |
| rs12981080 | 0.865825003 | 19 | 46117532 | T | 0.1797 |
| rs2015708 | 0.865825003 | 3 | 1.96E+08 | T | 0.1975 |
| rs35619957 | 0.865825003 | 2 | 1.26E+08 | A | 7.58E-05 |
| rs36155323 | 0.865825003 | 16 | 70694991 | G | 0.768 |
| rs73218870 | 0.865825003 | 21 | 21850181 | A | 0.4345 |
| rs7514343 | 0.865825003 | 1 | 41028394 | C | 0.2306 |
| rs79638902 | 0.865825003 | 19 | 45221767 | T | 0.9389 |
| rs11704423 | 0.867310023 | 22 | 23641261 | A | 1.30E-05 |
| rs9891876 | 0.867310023 | 17 | 71741401 | T | 0.7985 |
| rs35677603 | 0.869969983 | 11 | 47268310 | T | 0.1837 |
| rs6533603 | 0.869969983 | 4 | 1.13E+08 | T | 0.9117 |
| rs7557129 | 0.869969983 | 2 | 1.35E+08 | G | 0.3085 |
| rs7794532 | 0.869969983 | 7 | 1E+08 | A | 0.3055 |
| rs9268129 | 0.869969983 | 6 | 32254225 | C | 0.1484 |
| rs4468734 | 0.870118551 | 19 | 45086946 | A | 0.1387 |
| rs10400902 | 0.871485772 | 15 | 79231616 | A | 0.06849 |
| rs11651769 | 0.871485772 | 17 | 5162188 | G | 0.7049 |
| rs11825598 | 0.871485772 | 11 | 85634734 | C | 0.3216 |
| rs12122809 | 0.871485772 | 1 | 2.01E+08 | T | 0.9837 |
| rs2098088 | 0.871485772 | 19 | 45799975 | G | 0.5362 |
| rs34978823 | 0.871485772 | 3 | 57018247 | T | 0.3839 |
| rs519113 | 0.871485772 | 19 | 45376284 | G | 0.08926 |
| rs56059847 | 0.871485772 | 2 | 37476688 | G | 0.9171 |
| rs61977322 | 0.871485772 | 14 | 92957176 | C | 0.0561 |
| rs7597763 | 0.871485772 | 2 | 2.34E+08 | C | 0.5355 |
| rs76847167 | 0.871485772 | 9 | 30249215 | A | 0.8458 |
| rs7748777 | 0.871485772 | 6 | 41133806 | A | 0.8682 |
| rs17800789 | 0.88211724 | 19 | 45015020 | C | 0.03489 |
| rs2189966 | 0.88211724 | 7 | 28172066 | C | 0.2547 |
| rs28468656 | 0.88211724 | 9 | 95849625 | G | 0.1507 |
| rs12984266 | 0.882712839 | 19 | 45193534 | A | 0.07631 |
| rs12225345 | 0.893263121 | 11 | 65647105 | G | 0.6397 |
| rs35832098 | 0.893263121 | 4 | 1.1E+08 | A | 0.275 |
| rs7241845 | 0.893263121 | 18 | 2394400 | T | 8.92E-05 |
| rs8009153 | 0.893263121 | 14 | 92850461 | T | 0.913 |
| rs8109472 | 0.893281484 | 19 | 45736003 | C | 0.378 |
| rs12528892 | 0.899903152 | 6 | 32693506 | T | 0.5819 |
| rs150575877 | 0.899903152 | 19 | 46056134 | T | 0.4344 |
| 17:5008236:C:T | 0.906466518 | 17 | 5008236 | T | 0.4885 |
| rs111827758 | 0.906466518 | 15 | 59250404 | C | 0.05431 |
| rs1151103 | 0.906466518 | 11 | 59836391 | A | 0.198 |
| rs12775090 | 0.906466518 | 10 | 7450208 | A | 0.1371 |
| rs157902 | 0.906466518 | 7 | 1.31E+08 | A | 0.4592 |
| rs176474 | 0.906466518 | 7 | 1.06E+08 | C | 0.8925 |
| rs1884913 | 0.906466518 | 20 | 54984064 | C | 0.8791 |
| rs2341635 | 0.906466518 | 19 | 46470434 | A | 0.328 |
| rs2389488 | 0.906466518 | 4 | 1.19E+08 | G | 0.2599 |
| rs268120 | 0.906466518 | 2 | 65604914 | A | 0.2406 |
| rs2965162 | 0.906466518 | 19 | 45198936 | T | 0.4903 |
| rs3178166 | 0.906466518 | 19 | 45594170 | G | 0.4842 |
| rs335473 | 0.906466518 | 5 | 1.77E+08 | A | 0.06091 |
| rs34374273 | 0.906466518 | 19 | 45338895 | A | 0.4204 |
| rs34565850 | 0.906466518 | 11 | 85836356 | A | 0.04927 |
| rs346763 | 0.906466518 | 19 | 45729275 | A | 0.114 |
| rs3752229 | 0.906466518 | 19 | 1041352 | G | 0.08164 |
| rs4807455 | 0.906466518 | 19 | 1033559 | A | 0.6341 |
| rs605003 | 0.906466518 | 19 | 45708239 | G | 0.2883 |
| rs62044385 | 0.906466518 | 16 | 16907324 | C | 0.7622 |
| rs6455537 | 0.906466518 | 6 | 1.69E+08 | C | 4.43E-05 |
| rs646260 | 0.906466518 | 11 | 85824288 | G | 0.2116 |
| rs6714826 | 0.906466518 | 2 | 86777825 | C | 0.4272 |
| rs6983452 | 0.906466518 | 8 | 27448028 | T | 0.9818 |
| rs7272308 | 0.906466518 | 20 | 48583726 | A | 0.6136 |
| rs7559 | 0.906466518 | 3 | 1.87E+08 | C | 0.6842 |
| rs9276627 | 0.906466518 | 6 | 32743835 | T | 0.712 |
| rs9908652 | 0.906466518 | 17 | 2309117 | A | 0.2704 |
| 1:207653364:A:G | 0.906609424 | 1 | 2.08E+08 | G | 0.44 |
| rs9331916 | 0.907869187 | 8 | 27462150 | T | 0.4021 |
| rs5754508 | 0.91038088 | 22 | 21999229 | G | 0.9625 |
| rs11086109 | 0.916802366 | 19 | 18511230 | C | 0.5865 |
| rs17714718 | 0.916802366 | 19 | 45050747 | C | 0.8285 |
| rs2276044 | 0.916802366 | 11 | 59814287 | G | 0.9806 |
| rs4731 | 0.916802366 | 8 | 11666337 | G | 0.05479 |
| rs9586317 | 0.916802366 | 13 | 1.05E+08 | A | 0.7885 |
| 19:45200634:C:T | 0.920476192 | 19 | 45200634 | T | 0.5946 |
| 6:40997833:G:T | 0.920476192 | 6 | 40997833 | T | 0.6915 |
| rs73041776 | 0.920476192 | 19 | 33372243 | T | 0.3203 |
| rs8056564 | 0.920476192 | 16 | 81980019 | T | 0.0264 |
| 19:45429708:G:C | 0.92122618 | 19 | 45429708 | C | 0.1933 |
| rs10792828 | 0.92122618 | 11 | 85826797 | G | 0.4113 |
| rs11153444 | 0.92122618 | 6 | 1.14E+08 | C | 9.82E-05 |
| rs62118211 | 0.92122618 | 19 | 45103149 | A | 0.7572 |
| rs112659010 | 0.921808989 | 18 | 29562795 | C | 0.4353 |
| rs11635107 | 0.924933179 | 15 | 80414718 | C | 8.43E-05 |
| rs356181 | 0.924933179 | 4 | 90626139 | G | 0.5097 |
| rs3935067 | 0.924933179 | 7 | 1.43E+08 | C | 0.526 |
| rs4704850 | 0.924933179 | 5 | 1.57E+08 | A | 0.6693 |
| rs511104 | 0.924933179 | 1 | 2.01E+08 | G | 0.9987 |
| rs61932117 | 0.924933179 | 12 | 1.14E+08 | G | 0.8782 |
| rs6668174 | 0.927855032 | 1 | 1.61E+08 | C | 0.1891 |
| rs17218817 | 0.928242045 | 4 | 11038268 | G | 0.9762 |
| rs8108277 | 0.93666577 | 19 | 45205877 | C | 0.3684 |
| rs9401134 | 0.93666577 | 6 | 98199541 | C | 0.9308 |
| rs28394864 | 0.936723176 | 17 | 47450775 | A | 0.3782 |
| rs11532670 | 0.940586986 | 7 | 37371598 | G | 4.31E-05 |
| rs7084953 | 0.940586986 | 10 | 61755297 | T | 0.3116 |
| rs60721162 | 0.940604732 | 19 | 341500 | G | 0.9283 |
| 8:51466904:C:G | 0.941760535 | 8 | 51466904 | C | 1.45E-05 |
| rs12920716 | 0.941760535 | 16 | 81806977 | G | 0.8043 |
| 17:42597539:C:G | 0.942742395 | 17 | 42597539 | G | 0.9243 |
| 17:47293329:T:C | 0.942742395 | 17 | 47293329 | C | 0.2557 |
| 18:24241671:G:T | 0.942742395 | 18 | 24241671 | T | 0.4371 |
| 19:45325391:C:T | 0.942742395 | 19 | 45325391 | T | 0.364 |
| 1:21220674:T:G | 0.942742395 | 1 | 21220674 | T | 0.03166 |
| 1:4539246:T:C | 0.942742395 | 1 | 4539246 | C | 2.35E-05 |
| 21:27534261:T:C | 0.942742395 | 21 | 27534261 | C | 0.1887 |
| 2:127893897:G:C | 0.942742395 | 2 | 1.28E+08 | C | 0.269 |
| 3:158684240:C:T | 0.942742395 | 3 | 1.59E+08 | T | 9.56E-05 |
| 3:57705131:C:G | 0.942742395 | 3 | 57705131 | C | 0.8196 |
| 5:79582522:A:T | 0.942742395 | 5 | 79582522 | T | 0.03931 |
| 7:100042413:T:C | 0.942742395 | 7 | 1E+08 | T | 0.852 |
| 7:37877013:T:C | 0.942742395 | 7 | 37877013 | T | 0.6116 |
| 9:20579082:C:T | 0.942742395 | 9 | 20579082 | C | 0.6611 |
| rs10416426 | 0.942742395 | 19 | 33504177 | C | 0.3259 |
| rs10421035 | 0.942742395 | 19 | 45356674 | G | 0.9737 |
| rs1065712 | 0.942742395 | 8 | 11702122 | C | 0.4737 |
| rs10950558 | 0.942742395 | 7 | 15336391 | G | 6.61E-05 |
| rs10992628 | 0.942742395 | 9 | 95854235 | T | 0.1131 |
| rs111677930 | 0.942742395 | 1 | 6989416 | G | 0.5628 |
| rs112138840 | 0.942742395 | 14 | 85187781 | C | 0.9642 |
| rs11234562 | 0.942742395 | 11 | 85863769 | G | 0.9753 |
| rs113426708 | 0.942742395 | 17 | 4823657 | A | 0.7318 |
| rs11669351 | 0.942742395 | 19 | 1001100 | A | 0.6869 |
| rs11669713 | 0.942742395 | 19 | 44461049 | T | 0.2794 |
| rs1171830 | 0.942742395 | 10 | 61665886 | A | 0.2759 |
| rs12185072 | 0.942742395 | 15 | 58683410 | T | 7.92E-07 |
| rs12606312 | 0.942742395 | 18 | 28768448 | A | 0.5649 |
| rs12801188 | 0.942742395 | 11 | 47388214 | A | 0.1958 |
| rs12878801 | 0.942742395 | 14 | 92935944 | T | 0.9702 |
| rs17125944 | 0.942742395 | 14 | 53400629 | C | 0.04872 |
| rs17188860 | 0.942742395 | 1 | 2.11E+08 | A | 3.86E-05 |
| rs17236194 | 0.942742395 | 15 | 59002755 | C | 0.8035 |
| rs2327832 | 0.942742395 | 6 | 1.38E+08 | G | 0.402 |
| rs2966698 | 0.942742395 | 7 | 1.43E+08 | T | 0.8398 |
| rs3129882 | 0.942742395 | 6 | 32409530 | G | 0.7015 |
| rs34294852 | 0.942742395 | 5 | 1.5E+08 | C | 0.8529 |
| rs34346157 | 0.942742395 | 6 | 41166310 | A | 0.3348 |
| rs35385129 | 0.942742395 | 19 | 45162189 | A | 0.7957 |
| rs3760842 | 0.942742395 | 19 | 46217026 | A | 0.6889 |
| rs3820759 | 0.942742395 | 2 | 1.28E+08 | G | 0.9271 |
| rs4243226 | 0.942742395 | 16 | 81993974 | A | 0.07075 |
| rs4802053 | 0.942742395 | 19 | 40147239 | A | 0.7878 |
| rs4887131 | 0.942742395 | 15 | 74598757 | A | 0.4454 |
| rs56394238 | 0.942742395 | 19 | 45335676 | G | 0.3295 |
| rs62502392 | 0.942742395 | 8 | 27190945 | A | 0.2312 |
| rs6743562 | 0.942742395 | 2 | 50649362 | C | 5.20E-05 |
| rs67938966 | 0.942742395 | 9 | 8078979 | T | 0.1854 |
| rs6985003 | 0.942742395 | 8 | 1.45E+08 | T | 0.6203 |
| rs6996116 | 0.942742395 | 8 | 36382695 | G | 0.3092 |
| rs71488451 | 0.942742395 | 11 | 60035618 | G | 0.859 |
| rs72924626 | 0.942742395 | 11 | 60095740 | C | 0.1113 |
| rs73034893 | 0.942742395 | 19 | 45724044 | T | 0.7692 |
| rs74172479 | 0.942742395 | 19 | 45074764 | A | 0.6096 |
| rs7555072 | 0.942742395 | 1 | 90501379 | C | 6.47E-05 |
| rs7785860 | 0.942742395 | 7 | 12103271 | C | 0.345 |
| rs79451156 | 0.942742395 | 8 | 17975346 | G | 0.6065 |
| rs79627176 | 0.942742395 | 1 | 17922600 | G | 5.87E-06 |
| rs8015234 | 0.942742395 | 14 | 92851708 | A | 0.5465 |
| rs8112940 | 0.942742395 | 19 | 33345945 | A | 0.7858 |
| rs9268528 | 0.942742395 | 6 | 32383108 | G | 0.3372 |
| rs9448565 | 0.942742395 | 6 | 79486518 | T | 7.20E-05 |
| rs9581791 | 0.942742395 | 13 | 27547341 | A | 4.32E-05 |
| rs962607 | 0.942742395 | 6 | 1.14E+08 | T | 0.7475 |
| rs56035750 | 0.944769143 | 1 | 51496464 | A | 0.06111 |
| rs78777228 | 0.944769143 | 19 | 45963927 | G | 0.9653 |
| 19:54782407:C:T | 0.945215956 | 19 | 54782407 | C | 0.2164 |
| rs1081105 | 0.945215956 | 19 | 45412955 | C | 0.01363 |
| rs12894551 | 0.945215956 | 14 | 92922366 | T | 0.6352 |
| rs12907724 | 0.945215956 | 15 | 58916392 | G | 0.3746 |
| rs16893889 | 0.945215956 | 6 | 28237172 | G | 0.9594 |
| rs3134996 | 0.945215956 | 6 | 32636866 | A | 0.5512 |
| rs72843213 | 0.945215956 | 6 | 32299173 | T | 0.2214 |
| rs849877 | 0.945215956 | 6 | 22298699 | C | 0.4858 |
| rs10091076 | 0.947594458 | 8 | 1.04E+08 | C | 0.9367 |
| rs2318110 | 0.947594458 | 11 | 63700443 | G | 0.5498 |
| 8:27460368:G:C | 0.948299093 | 8 | 27460368 | C | 0.1656 |
| rs8076152 | 0.948299093 | 17 | 43995932 | G | 0.601 |
| 2:36936091:G:C | 0.949417613 | 2 | 36936091 | C | 0.1217 |
| 16:79588064:C:T | 0.951445518 | 16 | 79588064 | T | 0.5404 |
| 19:45215711:C:G | 0.951445518 | 19 | 45215711 | G | 0.09111 |
| 2:127881368:A:G | 0.951445518 | 2 | 1.28E+08 | A | 0.8592 |
| rs10454495 | 0.951445518 | 11 | 85895909 | G | 0.9409 |
| rs11731929 | 0.951445518 | 4 | 1.37E+08 | A | 0.5853 |
| rs11736866 | 0.951445518 | 4 | 831181 | A | 0.9548 |
| rs12589503 | 0.951445518 | 14 | 92894774 | A | 0.6187 |
| rs12739089 | 0.951445518 | 1 | 1.61E+08 | C | 0.04972 |
| rs412776 | 0.951445518 | 19 | 45379516 | A | 0.03318 |
| rs4904917 | 0.951445518 | 14 | 92927201 | C | 0.09636 |
| rs116462788 | 0.954168237 | 5 | 1.57E+08 | C | 0.8585 |
| rs13200221 | 0.954168237 | 6 | 47494767 | A | 0.8337 |
| rs28617688 | 0.954168237 | 15 | 59082463 | C | 0.6239 |
| rs4251953 | 0.954168237 | 19 | 44151525 | T | 0.5719 |
| rs55682629 | 0.954168237 | 16 | 85173431 | G | 0.7376 |
| rs7544804 | 0.954168237 | 1 | 2.1E+08 | T | 0.3295 |
| rs13400939 | 0.955013145 | 2 | 1.28E+08 | A | 0.4583 |
| rs3997700 | 0.958056394 | 6 | 41219627 | A | 0.8006 |
| 19:45333834:T:C | 0.96020335 | 19 | 45333834 | T | 0.6819 |
| rs73030322 | 0.96020335 | 3 | 17700913 | C | 0.5122 |
| rs73404245 | 0.96020335 | 6 | 31410327 | A | 0.1889 |
| rs11150581 | 0.960270181 | 16 | 30030699 | C | 0.08904 |
| 19:1053524:C:G | 0.961197197 | 19 | 1053524 | G | 0.7341 |
| 6:32666863:G:T | 0.961197197 | 6 | 32666863 | T | 0.4423 |
| rs112360708 | 0.961197197 | 2 | 2.35E+08 | A | 0.5164 |
| rs13048498 | 0.961197197 | 21 | 43037361 | T | 0.37 |
| rs139290129 | 0.961197197 | 19 | 45587406 | C | 0.8634 |
| rs17762258 | 0.961197197 | 17 | 55903829 | G | 0.02688 |
| rs3740204 | 0.961197197 | 10 | 11505175 | G | 0.2719 |
| rs62472729 | 0.961197197 | 7 | 1.43E+08 | C | 0.1489 |
| rs7529425 | 0.961197197 | 1 | 1.61E+08 | A | 0.04352 |
| rs7800465 | 0.961197197 | 7 | 5257232 | G | 0.2008 |
| rs9367098 | 0.961197197 | 6 | 41198422 | A | 0.09211 |
| 6:32344575:G:T | 0.962575964 | 6 | 32344575 | T | 0.1751 |
| rs2624217 | 0.962575964 | 5 | 86445632 | G | 0.8206 |
| rs12148472 | 0.963861991 | 15 | 79231478 | C | 0.1202 |
| rs3925492 | 0.963861991 | 19 | 44919392 | A | 0.9415 |
| rs62375411 | 0.963861991 | 5 | 86327701 | G | 0.8006 |
| rs854793 | 0.965530322 | 17 | 18030524 | A | 0.4469 |
| 10:124973893:G:T | 0.966429434 | 10 | 1.25E+08 | T | 8.13E-05 |
| 10:63648804:A:G | 0.966429434 | 10 | 63648804 | G | 0.4212 |
| 11:33934972:G:T | 0.966429434 | 11 | 33934972 | G | 0.7336 |
| 15:63583159:G:C | 0.966429434 | 15 | 63583159 | G | 0.3389 |
| 17:34447091:T:C | 0.966429434 | 17 | 34447091 | C | 0.1495 |
| 17:47390014:G:C | 0.966429434 | 17 | 47390014 | C | 0.8425 |
| 19:45584692:C:T | 0.966429434 | 19 | 45584692 | T | 0.245 |
| 19:45981815:G:T | 0.966429434 | 19 | 45981815 | T | 0.9778 |
| 1:207750568:T:C | 0.966429434 | 1 | 2.08E+08 | T | 0.0538 |
| 1:21704239:C:T | 0.966429434 | 1 | 21704239 | T | 0.5711 |
| 2:127862472:G:C | 0.966429434 | 2 | 1.28E+08 | G | 0.6183 |
| 4:112909713:T:C | 0.966429434 | 4 | 1.13E+08 | C | 0.7136 |
| 6:22289132:C:T | 0.966429434 | 6 | 22289132 | T | 0.04121 |
| 7:99956290:C:G | 0.966429434 | 7 | 99956290 | G | 0.6033 |
| 9:6603111:T:C | 0.966429434 | 9 | 6603111 | C | 6.78E-05 |
| rs10401176 | 0.966429434 | 19 | 45253491 | T | 0.1781 |
| rs10408847 | 0.966429434 | 19 | 45634682 | C | 0.6169 |
| rs10413386 | 0.966429434 | 19 | 33404269 | T | 0.5725 |
| rs10416371 | 0.966429434 | 19 | 45660136 | C | 0.9527 |
| rs10419669 | 0.966429434 | 19 | 45298069 | A | 0.7172 |
| rs1042116 | 0.966429434 | 6 | 32798548 | A | 0.9537 |
| rs10458914 | 0.966429434 | 11 | 47836026 | A | 0.651 |
| rs1053504 | 0.966429434 | 4 | 40198842 | G | 0.678 |
| rs10736339 | 0.966429434 | 10 | 82059970 | G | 0.5989 |
| rs10761438 | 0.966429434 | 10 | 61780209 | A | 0.2604 |
| rs10953481 | 0.966429434 | 7 | 1.05E+08 | T | 0.3527 |
| rs11077855 | 0.966429434 | 17 | 74681753 | T | 2.15E-05 |
| rs111559044 | 0.966429434 | 11 | 1.08E+08 | C | 0.1897 |
| rs111844806 | 0.966429434 | 4 | 11013340 | A | 0.9121 |
| rs112366983 | 0.966429434 | 14 | 62517594 | A | 0.5268 |
| rs113706587 | 0.966429434 | 5 | 1.8E+08 | A | 0.3742 |
| rs115947447 | 0.966429434 | 5 | 1.57E+08 | T | 0.896 |
| rs11654510 | 0.966429434 | 17 | 74677362 | A | 8.13E-05 |
| rs11709534 | 0.966429434 | 3 | 1.19E+08 | A | 0.04751 |
| rs11879091 | 0.966429434 | 19 | 1082265 | C | 0.9662 |
| rs12058296 | 0.966429434 | 1 | 66402424 | A | 0.3101 |
| rs12147407 | 0.966429434 | 14 | 53265785 | A | 0.3625 |
| rs12197146 | 0.966429434 | 6 | 47409828 | C | 0.8916 |
| rs123187 | 0.966429434 | 19 | 45830947 | A | 0.5062 |
| rs12452642 | 0.966429434 | 17 | 53070357 | T | 0.3576 |
| rs12461027 | 0.966429434 | 19 | 45691126 | A | 0.6275 |
| rs12546944 | 0.966429434 | 8 | 1.26E+08 | A | 0.5687 |
| rs12968702 | 0.966429434 | 18 | 52487062 | C | 0.6733 |
| rs12981557 | 0.966429434 | 19 | 45969490 | C | 0.4745 |
| rs12986272 | 0.966429434 | 19 | 45894381 | A | 0.1117 |
| rs1460537 | 0.966429434 | 18 | 41506017 | G | 9.23E-05 |
| rs17208153 | 0.966429434 | 6 | 32194595 | C | 0.2067 |
| rs17650302 | 0.966429434 | 7 | 12143138 | T | 0.6452 |
| rs17714676 | 0.966429434 | 19 | 45046730 | C | 0.7777 |
| rs1808648 | 0.966429434 | 5 | 1.72E+08 | A | 6.10E-05 |
| rs1866694 | 0.966429434 | 8 | 62172668 | A | 2.61E-05 |
| rs1980496 | 0.966429434 | 6 | 32340070 | T | 0.7929 |
| rs1981549 | 0.966429434 | 7 | 99541636 | G | 0.5286 |
| rs2014576 | 0.966429434 | 19 | 46269076 | A | 0.1904 |
| rs203878 | 0.966429434 | 6 | 28048996 | G | 0.3545 |
| rs208811 | 0.966429434 | 20 | 37497339 | A | 0.03042 |
| rs2340534 | 0.966429434 | 8 | 95992330 | A | 0.639 |
| rs2350962 | 0.966429434 | 17 | 17614097 | G | 0.028 |
| rs2458285 | 0.966429434 | 8 | 1.04E+08 | T | 0.2514 |
| rs2624213 | 0.966429434 | 5 | 86440947 | C | 0.4436 |
| rs2830068 | 0.966429434 | 21 | 27496667 | T | 0.347 |
| rs283810 | 0.966429434 | 19 | 45388241 | G | 0.2225 |
| rs28573295 | 0.966429434 | 4 | 37130863 | G | 0.1984 |
| rs2868099 | 0.966429434 | 11 | 59988193 | A | 0.1798 |
| rs2869771 | 0.966429434 | 4 | 10968018 | C | 0.7861 |
| rs2929414 | 0.966429434 | 3 | 20075645 | C | 1.15E-05 |
| rs2957061 | 0.966429434 | 8 | 1676796 | A | 0.2289 |
| rs2965169 | 0.966429434 | 19 | 45251156 | C | 0.01602 |
| rs2992391 | 0.966429434 | 13 | 29831521 | C | 4.03E-05 |
| rs3212930 | 0.966429434 | 19 | 45927610 | G | 0.2481 |
| rs34546629 | 0.966429434 | 22 | 39802495 | C | 0.1036 |
| rs34587452 | 0.966429434 | 4 | 1009900 | C | 0.1064 |
| rs34722237 | 0.966429434 | 17 | 65143651 | T | 0.7983 |
| rs356176 | 0.966429434 | 4 | 90630801 | G | 0.9412 |
| rs35646539 | 0.966429434 | 19 | 1083532 | T | 0.6679 |
| rs35810109 | 0.966429434 | 10 | 1.29E+08 | T | 1.80E-05 |
| rs35905922 | 0.966429434 | 11 | 60080733 | A | 0.26 |
| rs35990442 | 0.966429434 | 6 | 31184949 | A | 0.7635 |
| rs3844143 | 0.966429434 | 11 | 85850243 | C | 0.02692 |
| rs3852865 | 0.966429434 | 19 | 51714065 | A | 0.5194 |
| rs3865444 | 0.966429434 | 19 | 51727962 | A | 0.9226 |
| rs393584 | 0.966429434 | 19 | 45377334 | A | 0.4411 |
| rs3936112 | 0.966429434 | 16 | 81970938 | T | 0.3109 |
| rs4421019 | 0.966429434 | 4 | 40309851 | A | 0.4605 |
| rs4455662 | 0.966429434 | 6 | 28340529 | G | 0.4992 |
| rs4502845 | 0.966429434 | 5 | 1.68E+08 | G | 0.3336 |
| rs4550514 | 0.966429434 | 17 | 61583856 | C | 0.2902 |
| rs4663105 | 0.966429434 | 2 | 1.28E+08 | C | 0.4694 |
| rs4803836 | 0.966429434 | 19 | 46137818 | A | 0.8143 |
| rs4834237 | 0.966429434 | 4 | 1.13E+08 | T | 0.7612 |
| rs4985369 | 0.966429434 | 16 | 70860468 | G | 0.3741 |
| rs4987082 | 0.966429434 | 17 | 47481374 | C | 0.7825 |
| rs50871 | 0.966429434 | 19 | 45862515 | C | 0.3524 |
| rs527162 | 0.966429434 | 11 | 85715736 | C | 0.3124 |
| rs55661415 | 0.966429434 | 11 | 59952316 | G | 0.6835 |
| rs560442 | 0.966429434 | 11 | 82426758 | T | 0.3278 |
| rs56260001 | 0.966429434 | 18 | 47789989 | G | 7.62E-05 |
| rs56364110 | 0.966429434 | 19 | 41029820 | T | 0.4494 |
| rs57355367 | 0.966429434 | 19 | 45698085 | G | 0.4648 |
| rs58128433 | 0.966429434 | 4 | 1.13E+08 | T | 0.7016 |
| rs5848 | 0.966429434 | 17 | 42430244 | T | 0.3562 |
| rs58643066 | 0.966429434 | 10 | 61545182 | G | 0.2146 |
| rs58817628 | 0.966429434 | 1 | 2.08E+08 | G | 0.1737 |
| rs60073596 | 0.966429434 | 17 | 17870974 | T | 0.4559 |
| rs61901745 | 0.966429434 | 11 | 59962781 | G | 0.1684 |
| rs62116896 | 0.966429434 | 19 | 45035074 | T | 0.3554 |
| rs636772 | 0.966429434 | 11 | 85724572 | A | 0.03055 |
| rs6431223 | 0.966429434 | 2 | 1.28E+08 | A | 0.4549 |
| rs6457681 | 0.966429434 | 6 | 32773497 | T | 0.7628 |
| rs6499775 | 0.966429434 | 16 | 55759867 | G | 0.09264 |
| rs6502844 | 0.966429434 | 17 | 5019668 | C | 0.8185 |
| rs6751833 | 0.966429434 | 2 | 1.36E+08 | T | 0.9406 |
| rs67667588 | 0.966429434 | 17 | 52829761 | C | 9.87E-05 |
| rs67752778 | 0.966429434 | 4 | 11032923 | A | 0.9602 |
| rs6799302 | 0.966429434 | 3 | 1.57E+08 | A | 0.09215 |
| rs6978679 | 0.966429434 | 7 | 1.06E+08 | G | 0.8861 |
| rs6981191 | 0.966429434 | 8 | 1.27E+08 | T | 0.1388 |
| rs7085411 | 0.966429434 | 10 | 11699441 | T | 0.1608 |
| rs7149030 | 0.966429434 | 14 | 92919504 | G | 0.2405 |
| rs7167430 | 0.966429434 | 15 | 33781323 | C | 5.95E-05 |
| rs7219226 | 0.966429434 | 17 | 47289566 | C | 0.4149 |
| rs7256075 | 0.966429434 | 19 | 1030956 | G | 0.4509 |
| rs72745053 | 0.966429434 | 15 | 59159259 | G | 0.7723 |
| rs72838225 | 0.966429434 | 2 | 1.28E+08 | T | 0.9285 |
| rs72874964 | 0.966429434 | 18 | 20764273 | A | 0.1297 |
| rs72893359 | 0.966429434 | 11 | 45560676 | G | 1.54E-05 |
| rs73035597 | 0.966429434 | 19 | 45199524 | T | 0.6971 |
| rs731170 | 0.966429434 | 19 | 55176262 | A | 0.8007 |
| rs7384878 | 0.966429434 | 7 | 99932049 | C | 0.8257 |
| rs73904091 | 0.966429434 | 20 | 31553023 | T | 0.4533 |
| rs7404856 | 0.966429434 | 16 | 77130339 | A | 1.94E-05 |
| rs752780 | 0.966429434 | 2 | 1.28E+08 | C | 0.544 |
| rs754894 | 0.966429434 | 1 | 2.03E+08 | C | 0.2212 |
| rs7555508 | 0.966429434 | 1 | 1.62E+08 | G | 4.93E-05 |
| rs76692773 | 0.966429434 | 19 | 45394211 | T | 0.8948 |
| rs77210365 | 0.966429434 | 5 | 80894657 | C | 0.04907 |
| rs7795740 | 0.966429434 | 7 | 88551357 | T | 3.77E-05 |
| rs78116022 | 0.966429434 | 1 | 65072074 | A | 0.2189 |
| rs79029793 | 0.966429434 | 16 | 90023272 | G | 0.3917 |
| rs79527490 | 0.966429434 | 2 | 1.28E+08 | T | 0.2052 |
| rs8005311 | 0.966429434 | 14 | 36466531 | G | 0.1581 |
| rs8021135 | 0.966429434 | 14 | 53450648 | G | 0.4195 |
| rs8100672 | 0.966429434 | 19 | 1021637 | T | 0.5487 |
| rs922447 | 0.966429434 | 16 | 84568007 | C | 0.9699 |
| rs9267951 | 0.966429434 | 6 | 32212655 | T | 0.5124 |
| rs9395288 | 0.966429434 | 6 | 47598125 | T | 0.156 |
| rs941287 | 0.966429434 | 7 | 99807473 | T | 0.2324 |
| rs9463349 | 0.966429434 | 6 | 47708815 | A | 0.4406 |
| rs9601273 | 0.966429434 | 13 | 80413817 | A | 0.9842 |
| rs9912738 | 0.966429434 | 17 | 47510584 | C | 0.5823 |
| rs2967668 | 0.967950673 | 19 | 45302951 | G | 0.1002 |
| rs12433380 | 0.968220744 | 14 | 93012967 | C | 0.7706 |
| rs138537907 | 0.968220744 | 19 | 1076681 | G | 0.568 |
| rs3789576 | 0.968220744 | 1 | 50945526 | T | 0.5521 |
| rs11659193 | 0.969422981 | 18 | 48830075 | C | 0.2023 |
| rs13230744 | 0.969422981 | 7 | 99797792 | G | 0.1676 |
| rs3783448 | 0.969422981 | 14 | 53176982 | C | 0.03808 |
| 11:47547046:T:C | 0.969866645 | 11 | 47547046 | C | 0.02126 |
| rs1039405 | 0.969866645 | 19 | 51626237 | A | 0.8692 |
| rs11672923 | 0.969866645 | 19 | 45802022 | T | 0.8562 |
| rs56159792 | 0.969866645 | 10 | 15313310 | G | 0.1885 |
| rs61268362 | 0.969866645 | 14 | 1.07E+08 | C | 0.7575 |
| 17:4974111:T:C | 0.970047457 | 17 | 4974111 | C | 0.5812 |
| 1:161186313:C:T | 0.970047457 | 1 | 1.61E+08 | T | 0.8379 |
| 21:28091723:A:T | 0.970047457 | 21 | 28091723 | T | 0.1636 |
| 4:89879212:C:T | 0.970047457 | 4 | 89879212 | C | 2.20E-05 |
| rs10897024 | 0.970047457 | 11 | 60055651 | A | 0.05415 |
| rs10945496 | 0.970047457 | 6 | 1.69E+08 | G | 3.92E-05 |
| rs112262807 | 0.970047457 | 19 | 45331686 | T | 0.4915 |
| rs11234568 | 0.970047457 | 11 | 85876222 | G | 0.03237 |
| rs11680911 | 0.970047457 | 2 | 1.28E+08 | C | 0.3545 |
| rs11771145 | 0.970047457 | 7 | 1.43E+08 | A | 0.2788 |
| rs12972306 | 0.970047457 | 19 | 45079474 | T | 0.1256 |
| rs12978931 | 0.970047457 | 19 | 45363700 | G | 0.2043 |
| rs1967311 | 0.970047457 | 19 | 45819307 | G | 0.9505 |
| rs199604 | 0.970047457 | 20 | 19971112 | A | 4.96E-05 |
| rs2004357 | 0.970047457 | 19 | 45618959 | A | 0.3825 |
| rs238406 | 0.970047457 | 19 | 45868309 | T | 0.5678 |
| rs2927437 | 0.970047457 | 19 | 45241638 | A | 0.9375 |
| rs396960 | 0.970047457 | 6 | 32191581 | A | 0.4046 |
| rs418891 | 0.970047457 | 17 | 43693538 | T | 0.3667 |
| rs4792945 | 0.970047457 | 17 | 42552832 | C | 0.4397 |
| rs536332 | 0.970047457 | 8 | 27475767 | G | 0.8602 |
| rs60220759 | 0.970047457 | 17 | 72814387 | G | 0.7587 |
| rs6064398 | 0.970047457 | 20 | 55046059 | G | 0.9731 |
| rs6910948 | 0.970047457 | 6 | 22306698 | C | 0.09719 |
| rs7257940 | 0.970047457 | 19 | 43876120 | A | 0.2777 |
| rs72974886 | 0.970047457 | 11 | 1.03E+08 | G | 3.29E-06 |
| rs73558194 | 0.970047457 | 19 | 45483844 | A | 0.7786 |
| rs7537669 | 0.970047457 | 1 | 2.08E+08 | C | 0.2918 |
| rs8113514 | 0.970047457 | 19 | 45693697 | G | 0.6204 |
| rs9683415 | 0.970047457 | 4 | 40291916 | A | 0.33 |
| rs12978800 | 0.970410968 | 19 | 45724633 | T | 0.8012 |
| 11:47380340:G:T | 0.97051262 | 11 | 47380340 | G | 0.1033 |
| 11:7211896:C:G | 0.97051262 | 11 | 7211896 | C | 0.5457 |
| 11:85871184:G:C | 0.97051262 | 11 | 85871184 | C | 0.06795 |
| 13:19729760:C:T | 0.97051262 | 13 | 19729760 | C | 7.85E-05 |
| 17:44246624:C:A | 0.97051262 | 17 | 44246624 | A | 0.3995 |
| 19:45080065:C:G | 0.97051262 | 19 | 45080065 | C | 0.7948 |
| 19:45354238:G:C | 0.97051262 | 19 | 45354238 | G | 0.6283 |
| 19:45448465:T:G | 0.97051262 | 19 | 45448465 | G | 0.03787 |
| 19:51723546:C:G | 0.97051262 | 19 | 51723546 | G | 0.9097 |
| 1:229222734:G:C | 0.97051262 | 1 | 2.29E+08 | C | 0.8821 |
| 21:40757973:T:G | 0.97051262 | 21 | 40757973 | T | 0.8001 |
| 3:154771786:G:C | 0.97051262 | 3 | 1.55E+08 | C | 0.9431 |
| 6:20092949:T:C | 0.97051262 | 6 | 20092949 | C | 0.4402 |
| 6:31329494:C:T | 0.97051262 | 6 | 31329494 | C | 0.7284 |
| 8:27217763:C:T | 0.97051262 | 8 | 27217763 | T | 0.3837 |
| 9:27221114:T:G | 0.97051262 | 9 | 27221114 | G | 0.9702 |
| rs10011598 | 0.97051262 | 4 | 31228516 | G | 2.80E-05 |
| rs1007369 | 0.97051262 | 11 | 1.2E+08 | C | 5.03E-05 |
| rs10142146 | 0.97051262 | 14 | 92888341 | T | 0.6733 |
| rs10254366 | 0.97051262 | 7 | 1.29E+08 | A | 0.6784 |
| rs1046545 | 0.97051262 | 3 | 57305170 | T | 0.725 |
| rs10863923 | 0.97051262 | 1 | 2.12E+08 | A | 0.1851 |
| rs10932008 | 0.97051262 | 2 | 2.04E+08 | G | 0.3776 |
| rs111278892 | 0.97051262 | 19 | 1039323 | G | 0.3912 |
| rs11137524 | 0.97051262 | 9 | 91766706 | A | 0.7238 |
| rs113681266 | 0.97051262 | 2 | 38662281 | G | 2.14E-05 |
| rs1143679 | 0.97051262 | 16 | 31276811 | A | 0.3169 |
| rs11754574 | 0.97051262 | 6 | 34794961 | A | 0.6868 |
| rs11881756 | 0.97051262 | 19 | 45220896 | C | 0.6959 |
| rs12151021 | 0.97051262 | 19 | 1050874 | A | 0.1518 |
| rs12459810 | 0.97051262 | 19 | 45249661 | T | 0.4692 |
| rs12461065 | 0.97051262 | 19 | 45605308 | T | 0.3857 |
| rs12525381 | 0.97051262 | 6 | 696327 | A | 0.7469 |
| rs12617835 | 0.97051262 | 2 | 1.28E+08 | C | 0.3064 |
| rs13136820 | 0.97051262 | 4 | 40307564 | C | 0.4158 |
| rs13269897 | 0.97051262 | 8 | 1.17E+08 | C | 1.50E-05 |
| rs146335962 | 0.97051262 | 19 | 47154484 | T | 0.1354 |
| rs1690543 | 0.97051262 | 1 | 83132585 | C | 0.1232 |
| rs17014773 | 0.97051262 | 2 | 1.28E+08 | G | 0.3414 |
| rs1799786 | 0.97051262 | 19 | 45868038 | A | 0.2137 |
| rs199498 | 0.97051262 | 17 | 44865603 | C | 0.8732 |
| rs1998098 | 0.97051262 | 14 | 92938265 | A | 0.8537 |
| rs2070902 | 0.97051262 | 1 | 1.61E+08 | T | 0.6037 |
| rs2241038 | 0.97051262 | 16 | 90088904 | G | 0.5386 |
| rs2338812 | 0.97051262 | 5 | 1.41E+08 | G | 0.2427 |
| rs2395516 | 0.97051262 | 6 | 32580657 | C | 0.6613 |
| rs2565067 | 0.97051262 | 8 | 27331119 | A | 0.939 |
| rs2613776 | 0.97051262 | 19 | 5076777 | T | 0.354 |
| rs2631307 | 0.97051262 | 17 | 42094638 | G | 0.9593 |
| rs2761421 | 0.97051262 | 1 | 2.08E+08 | C | 0.9663 |
| rs2830435 | 0.97051262 | 21 | 28092253 | G | 0.04699 |
| rs28393946 | 0.97051262 | 15 | 51047085 | A | 0.9692 |
| rs34181358 | 0.97051262 | 8 | 27322974 | A | 0.02004 |
| rs34311866 | 0.97051262 | 4 | 951947 | C | 0.924 |
| rs34457613 | 0.97051262 | 16 | 81982291 | G | 0.1184 |
| rs35103166 | 0.97051262 | 2 | 1.28E+08 | C | 0.3832 |
| rs35504048 | 0.97051262 | 13 | 81232288 | C | 0.2521 |
| rs35805829 | 0.97051262 | 11 | 47805478 | C | 0.02978 |
| rs3669 | 0.97051262 | 19 | 45323073 | C | 0.149 |
| rs3763313 | 0.97051262 | 6 | 32376471 | C | 0.9762 |
| rs3806157 | 0.97051262 | 6 | 32373801 | G | 0.7194 |
| rs4504245 | 0.97051262 | 4 | 11014822 | A | 0.9177 |
| rs4722755 | 0.97051262 | 7 | 28152836 | A | 0.4918 |
| rs4726618 | 0.97051262 | 7 | 1.43E+08 | T | 0.1389 |
| rs4745489 | 0.97051262 | 9 | 78661768 | A | 1.71E-05 |
| rs4873494 | 0.97051262 | 8 | 51963714 | G | 0.8812 |
| rs5158 | 0.97051262 | 19 | 45447178 | T | 0.5423 |
| rs55690962 | 0.97051262 | 11 | 85875634 | G | 0.8675 |
| rs55906136 | 0.97051262 | 5 | 1.54E+08 | A | 0.1727 |
| rs56140728 | 0.97051262 | 19 | 41136907 | T | 0.7497 |
| rs57719175 | 0.97051262 | 13 | 96175396 | A | 1.80E-05 |
| rs59120146 | 0.97051262 | 7 | 1E+08 | A | 0.7082 |
| rs6123522 | 0.97051262 | 20 | 36889021 | G | 3.70E-05 |
| rs62120565 | 0.97051262 | 19 | 45197027 | C | 0.4306 |
| rs6460906 | 0.97051262 | 7 | 12283276 | C | 0.649 |
| rs647775 | 0.97051262 | 10 | 34318258 | A | 9.31E-05 |
| rs6504163 | 0.97051262 | 17 | 61545779 | C | 0.5312 |
| rs655931 | 0.97051262 | 18 | 34907964 | A | 0.3182 |
| rs659018 | 0.97051262 | 11 | 85652688 | A | 0.3484 |
| rs663925 | 0.97051262 | 11 | 59815517 | G | 0.139 |
| rs664034 | 0.97051262 | 11 | 59930966 | C | 0.0638 |
| rs7248283 | 0.97051262 | 19 | 45208682 | T | 0.2584 |
| rs72814575 | 0.97051262 | 2 | 57661879 | C | 0.9797 |
| rs72971649 | 0.97051262 | 19 | 995918 | C | 0.77 |
| rs72973584 | 0.97051262 | 19 | 1046076 | T | 0.1567 |
| rs7300063 | 0.97051262 | 12 | 94646779 | C | 0.05641 |
| rs73050293 | 0.97051262 | 19 | 45379746 | G | 0.89 |
| rs7313875 | 0.97051262 | 12 | 67649137 | G | 0.005798 |
| rs73223431 | 0.97051262 | 8 | 27219987 | T | 0.7037 |
| rs73430140 | 0.97051262 | 12 | 1.14E+08 | A | 0.6564 |
| rs7470777 | 0.97051262 | 9 | 1.36E+08 | G | 0.6027 |
| rs75509651 | 0.97051262 | 19 | 45619506 | A | 0.09112 |
| rs7807156 | 0.97051262 | 7 | 99686116 | G | 0.01013 |
| rs7808752 | 0.97051262 | 7 | 1.29E+08 | A | 0.7764 |
| rs8108110 | 0.97051262 | 19 | 45234124 | A | 0.2029 |
| rs832505 | 0.97051262 | 12 | 94672735 | T | 0.3055 |
| rs897148 | 0.97051262 | 8 | 1.27E+08 | G | 0.5373 |
| rs9267525 | 0.97051262 | 6 | 31607942 | C | 0.1154 |
| rs9267542 | 0.97051262 | 6 | 31668049 | T | 0.5141 |
| rs9301411 | 0.97051262 | 13 | 1.1E+08 | G | 0.914 |
| rs9640386 | 0.97051262 | 7 | 1.43E+08 | G | 0.1111 |
| rs9759516 | 0.97051262 | 4 | 40291084 | C | 0.4319 |
| 2:4164087:G:T | 0.970788211 | 2 | 4164087 | T | 9.51E-05 |
| rs35188103 | 0.971027769 | 14 | 92921725 | T | 0.3223 |
| rs359564 | 0.971064758 | 3 | 1.55E+08 | T | 0.2988 |
| rs12273113 | 0.971688705 | 11 | 85759778 | A | 0.8655 |
| rs17617 | 0.971688705 | 1 | 2.08E+08 | C | 0.7247 |
| rs12151229 | 0.972343013 | 19 | 45848392 | T | 0.8751 |
| rs12950511 | 0.972343013 | 17 | 47320938 | T | 0.6834 |
| rs1426248 | 0.972343013 | 11 | 60047410 | A | 0.07893 |
| rs204480 | 0.972343013 | 19 | 45477111 | T | 0.05716 |
| rs253296 | 0.972343013 | 5 | 1.5E+08 | A | 0.6134 |
| rs296522 | 0.972343013 | 1 | 2.01E+08 | T | 0.1747 |
| rs317656 | 0.972343013 | 12 | 69681101 | T | 0.815 |
| rs35836101 | 0.972343013 | 19 | 45616982 | A | 0.3013 |
| rs67598967 | 0.972343013 | 11 | 85843197 | A | 0.3763 |
| rs6771887 | 0.972343013 | 3 | 1.36E+08 | A | 0.3076 |
| rs9328259 | 0.972343013 | 6 | 508972 | C | 0.3 |
| rs9530949 | 0.972343013 | 13 | 80302125 | C | 0.9752 |
| rs10788284 | 0.973109667 | 10 | 1.24E+08 | C | 0.4314 |
| rs4330 | 0.973109667 | 17 | 61563661 | C | 0.6561 |
| rs80299306 | 0.973109667 | 16 | 70718300 | G | 0.447 |
| rs9394754 | 0.973109667 | 6 | 41000432 | A | 0.2518 |
| rs72932709 | 0.975103881 | 2 | 2.04E+08 | G | 0.2257 |
| rs10401439 | 0.975315762 | 19 | 46320780 | T | 0.4546 |
| rs2097442 | 0.980057279 | 6 | 32422191 | A | 0.7232 |
| rs687938 | 0.980057279 | 4 | 82733327 | T | 0.5611 |
| 10:61575320:C:T | 0.980365548 | 10 | 61575320 | T | 0.2125 |
| 19:41089274:C:T | 0.980365548 | 19 | 41089274 | T | 0.7012 |
| 6:31321407:C:T | 0.980365548 | 6 | 31321407 | T | 0.4344 |
| rs12906745 | 0.980365548 | 15 | 96069268 | A | 5.01E-05 |
| rs147901416 | 0.980365548 | 19 | 45466792 | A | 0.2922 |
| rs150535 | 0.980365548 | 5 | 95039580 | T | 3.09E-05 |
| rs2858331 | 0.980365548 | 6 | 32681277 | G | 0.8019 |
| rs2880532 | 0.980365548 | 13 | 19660878 | G | 7.74E-05 |
| rs346758 | 0.980365548 | 19 | 45725739 | G | 0.09347 |
| rs4147904 | 0.980365548 | 19 | 1040765 | A | 0.1438 |
| rs439156 | 0.980365548 | 5 | 1.18E+08 | T | 0.9464 |
| rs4720591 | 0.980365548 | 7 | 47347089 | A | 0.8743 |
| rs4940749 | 0.980365548 | 18 | 56413217 | T | 0.5043 |
| rs62117205 | 0.980365548 | 19 | 45255266 | C | 0.2383 |
| rs75364523 | 0.980365548 | 17 | 4723121 | A | 0.7712 |
| rs77241309 | 0.980365548 | 19 | 45349177 | C | 0.8703 |
| rs79924756 | 0.980365548 | 19 | 44321973 | G | 0.7174 |
| rs8100197 | 0.980365548 | 19 | 45253582 | A | 0.08981 |
| rs8623 | 0.980365548 | 1 | 1.68E+08 | T | 0.5266 |
| rs9520594 | 0.980365548 | 13 | 1.08E+08 | G | 0.9218 |
| rs77790943 | 0.980936253 | 6 | 41070941 | A | 0.6802 |
| rs66867801 | 0.983208096 | 19 | 45457293 | T | 0.02299 |
| 12:72185157:C:T | 0.9833593 | 12 | 72185157 | T | 0.577 |
| 19:45196663:G:C | 0.9833593 | 19 | 45196663 | C | 0.5238 |
| 19:45304028:T:C | 0.9833593 | 19 | 45304028 | C | 0.534 |
| 6:32658933:C:G | 0.9833593 | 6 | 32658933 | G | 0.2451 |
| rs1060743 | 0.9833593 | 2 | 1.28E+08 | G | 0.1783 |
| rs10792832 | 0.9833593 | 11 | 85867875 | A | 0.1042 |
| rs112104416 | 0.9833593 | 17 | 75100359 | T | 8.05E-06 |
| rs116986273 | 0.9833593 | 10 | 18260144 | G | 0.9842 |
| rs11759347 | 0.9833593 | 6 | 41167097 | A | 0.916 |
| rs11994116 | 0.9833593 | 8 | 1.27E+08 | A | 0.8249 |
| rs12401322 | 0.9833593 | 1 | 1.61E+08 | A | 0.4971 |
| rs17014873 | 0.9833593 | 2 | 1.28E+08 | C | 0.03921 |
| rs2072561 | 0.9833593 | 19 | 46525867 | G | 0.4122 |
| rs2223592 | 0.9833593 | 6 | 40965206 | C | 0.2179 |
| rs2245950 | 0.9833593 | 6 | 29943866 | A | 0.7398 |
| rs2525556 | 0.9833593 | 7 | 99577744 | C | 0.1174 |
| rs2849233 | 0.9833593 | 18 | 48331553 | C | 0.1825 |
| rs34874378 | 0.9833593 | 19 | 45516881 | A | 0.2248 |
| rs4844579 | 0.9833593 | 1 | 2.07E+08 | T | 0.7827 |
| rs4926257 | 0.9833593 | 19 | 13393537 | T | 2.85E-05 |
| rs55988502 | 0.9833593 | 3 | 1.55E+08 | G | 0.3367 |
| rs58145248 | 0.9833593 | 8 | 17358746 | A | 1.25E-05 |
| rs58147322 | 0.9833593 | 17 | 1655605 | T | 0.5917 |
| rs58581483 | 0.9833593 | 1 | 3126741 | C | 0.1667 |
| rs62048654 | 0.9833593 | 16 | 84579106 | G | 0.7306 |
| rs620807 | 0.9833593 | 19 | 45706952 | G | 0.4713 |
| rs62248062 | 0.9833593 | 3 | 71809960 | G | 0.8301 |
| rs6705834 | 0.9833593 | 2 | 2.34E+08 | C | 0.7215 |
| rs6945902 | 0.9833593 | 7 | 12286409 | A | 0.1809 |
| rs7078873 | 0.9833593 | 10 | 61787587 | A | 0.2054 |
| rs7251911 | 0.9833593 | 19 | 45582402 | G | 0.04895 |
| rs72839527 | 0.9833593 | 17 | 5340319 | C | 0.09741 |
| rs732594 | 0.9833593 | 6 | 35206553 | A | 0.6415 |
| rs76981546 | 0.9833593 | 6 | 47889051 | C | 0.5476 |
| rs8100391 | 0.9833593 | 19 | 1010973 | A | 0.1012 |
| rs846881 | 0.9833593 | 19 | 45078553 | C | 0.1813 |
| rs893434 | 0.9833593 | 2 | 1.28E+08 | A | 0.8736 |
| rs9268852 | 0.9833593 | 6 | 32429594 | A | 0.9211 |
| rs9959607 | 0.9833593 | 18 | 49081262 | A | 4.13E-05 |
| rs2158412 | 0.984721502 | 7 | 1.22E+08 | G | 0.3668 |
| rs1275394 | 0.987833594 | 20 | 35066318 | T | 0.4092 |
| rs1672452 | 0.987833594 | 12 | 69670259 | T | 0.6912 |
| rs221834 | 0.987833594 | 7 | 1E+08 | G | 0.7367 |
| rs3805096 | 0.987833594 | 2 | 15430264 | C | 1.87E-05 |
| rs467021 | 0.987833594 | 21 | 27517077 | C | 0.1145 |
| rs517835 | 0.987833594 | 11 | 1.26E+08 | C | 7.46E-05 |
| rs62385163 | 0.987833594 | 5 | 1.4E+08 | G | 0.3264 |
| rs73918299 | 0.987833594 | 19 | 996665 | G | 0.5983 |
| rs76935275 | 0.987833594 | 10 | 1.24E+08 | G | 0.6375 |
| rs886249 | 0.987833594 | 17 | 43946291 | G | 0.4357 |
| rs12463319 | 0.989763214 | 19 | 44282857 | T | 0.1843 |
| rs11257180 | 0.990807705 | 10 | 11623983 | C | 0.585 |
| rs34042178 | 0.990807705 | 7 | 1.24E+08 | C | 5.40E-05 |
| rs4323219 | 0.990807705 | 5 | 1.56E+08 | G | 0.8044 |
| rs59960564 | 0.990807705 | 19 | 45216528 | G | 0.4488 |
| rs10936164 | 0.992428199 | 3 | 1.59E+08 | C | 7.35E-05 |
| rs1661176 | 0.992428199 | 19 | 45048379 | T | 0.7952 |
| rs321666 | 0.992428199 | 18 | 8631904 | A | 0.3603 |
| rs4900122 | 0.992428199 | 14 | 92905817 | A | 0.6779 |
| rs55715159 | 0.992428199 | 11 | 60045958 | C | 0.9626 |
| rs58816127 | 0.992428199 | 8 | 48996302 | G | 0.4576 |
| rs7809394 | 0.992428199 | 7 | 55145382 | T | 0.1575 |
| rs8108847 | 0.992428199 | 19 | 1015961 | G | 0.02701 |
| rs10194375 | 0.993849962 | 2 | 1.28E+08 | A | 0.1515 |
| rs2242660 | 0.993849962 | 6 | 31597753 | A | 0.8103 |
| rs344819 | 0.993849962 | 19 | 45823174 | G | 0.2872 |
| rs56089021 | 0.993849962 | 19 | 45629166 | A | 0.7663 |
| rs7085274 | 0.993849962 | 10 | 61785671 | A | 0.3166 |
| rs72920853 | 0.993849962 | 11 | 59998720 | A | 0.2065 |
| rs9268433 | 0.993849962 | 6 | 32345891 | G | 0.3817 |
| rs115710528 | 0.995376244 | 3 | 29285684 | G | 0.4973 |
| rs11845652 | 0.995376244 | 14 | 92875786 | G | 0.5456 |
| rs314326 | 0.995376244 | 7 | 1E+08 | C | 0.2126 |
| rs56402156 | 0.995376244 | 7 | 1.43E+08 | A | 0.7307 |
| 11:13864260:C:T | 0.998618604 | 11 | 13864260 | T | 5.51E-05 |
| 11:59854313:C:T | 0.998618604 | 11 | 59854313 | T | 0.9268 |
| 17:17954535:A:G | 0.998618604 | 17 | 17954535 | A | 0.4459 |
| 8:27560651:C:T | 0.998618604 | 8 | 27560651 | T | 0.473 |
| rs10109884 | 0.998618604 | 8 | 8299608 | G | 0.1906 |
| rs11074275 | 0.998618604 | 15 | 94957502 | A | 7.11E-05 |
| rs12442145 | 0.998618604 | 15 | 51013595 | A | 0.1288 |
| rs1292635 | 0.998618604 | 16 | 19842947 | A | 0.03853 |
| rs12989701 | 0.998618604 | 2 | 1.28E+08 | A | 0.1681 |
| rs13070907 | 0.998618604 | 3 | 1.96E+08 | A | 0.6596 |
| rs1731393 | 0.998618604 | 3 | 1.45E+08 | C | 8.98E-05 |
| rs187956549 | 0.998618604 | 12 | 67643018 | A | 0.8387 |
| rs2422599 | 0.998618604 | 20 | 1500480 | A | 0.3707 |
| rs2553022 | 0.998618604 | 7 | 1E+08 | G | 0.4847 |
| rs2584662 | 0.998618604 | 17 | 47470487 | C | 0.9764 |
| rs3129867 | 0.998618604 | 6 | 32404220 | G | 0.873 |
| rs34370781 | 0.998618604 | 1 | 1.61E+08 | C | 0.6246 |
| rs34644948 | 0.998618604 | 16 | 70681658 | T | 0.6434 |
| rs56073911 | 0.998618604 | 8 | 27298409 | A | 0.7704 |
| rs6014724 | 0.998618604 | 20 | 54998544 | G | 0.2832 |
| rs60967546 | 0.998618604 | 19 | 6873507 | C | 0.1283 |
| rs62489295 | 0.998618604 | 7 | 1.29E+08 | A | 0.372 |
| rs71533426 | 0.998618604 | 7 | 12220605 | A | 0.6188 |
| rs7249244 | 0.998618604 | 19 | 45245698 | C | 0.1643 |
| rs7799105 | 0.998618604 | 7 | 1.43E+08 | A | 0.0981 |
| rs78015303 | 0.998618604 | 17 | 5126768 | A | 0.8436 |
| rs905149 | 0.998618604 | 19 | 1066917 | G | 0.01143 |
| rs7412 | 0.999711332 | 19 | 45412079 | T | 0.8496 |

**Supplementary Table 6.** Welch Two Sample T-test Comparing Risk SNP Odds Ratio in risk/resilience (RS) SNP-pairs and risk/null (RN) SNP-pairs

data: snp_info$OR_risk[snp_info$SNP_Group == "RS"] and snp_info$OR_risk[snp_info$SNP_Group == "RN"]

t = -10.654, df = 161.13, p-value < 2.2e-16

alternative hypothesis: true difference in means is not equal to 0

95 percent confidence interval:

-0.2035089 -0.1398649

sample estimates:

mean of x mean of y

1.095441 1.267128

**Supplementary Table 7.** Welch Two Sample T-test Comparing Resilience SNP Odds Ratio in risk/resilience (RS) SNP-pairs and inverse-risk/null (BN) SNP-pairs

data: snp_info$OR_res[snp_info$SNP_Group == "RS"] and snp_info$OR_risk[snp_info$SNP_Group == "BN"]

t = 9.4948, df = 309.82, p-value < 2.2e-16

alternative hypothesis: true difference in means is not equal to 0

95 percent confidence interval:

0.05948311 0.09058169

sample estimates:

mean of x mean of y

0.8963404 0.8213080

| BB | 1 | s^2^r | s^2^r^2^ |
| --- | --- | --- | --- |
| Bb | 1 | sr | sr^2^ |
| bb | 1 | r | r^2^ |
|  | aa | Aa | AA |
| **Supplementary Table 8: 2-locus risk/resilience calculations**. The formulae used to calculate total risk for each simulated SNP pair, where ‘a’ and ‘A’ represent the major and minor allele for the risk SNP, ‘b’ and ‘B’ represent the major and minor alleles for the resilience SNP, s represents the odds ratio of resilience effects and r represents the odds ratio of risk effects. | | | |
