## Supplementary Methods for "A Novel Method to Disentangle Tightly Linked Risk and Resilience Genes for Brain Disorders: Application to Alzheimer’s Disease"

**SNP-Pair Simulation**

One million simulated individuals were randomly assigned to be either a case with a simulated polygenic disease or an unaffected control. In these individuals we simulated 4 types of SNP-pairs defined as two SNPs, with 2 alleles each, simulated to be in LD with an r^2^ between 0.5 and 1. The four pair types were: 1) risk/null pairs with one risk-associated SNP that was more common in cases and one null SNP that is unevenly distributed in cases and controls only through its LD with the risk SNP; 2) Inverse-risk/null pairs with one SNP that was *less* common in cases and one null SNP; 3) Risk/resilience pairs with one SNP that was more common in cases and one SNP that was less common in cases *and* which reduced the effects of the risk SNP when both were present; and 4) Null/null pairs with two SNPs in LD that were equally distributed in cases and controls. We randomly generated SNP-pairs weighted by predefined probabilities with risk/null, inverse-risk/null, and risk/resilience pairs equally frequent and null/null SNPs 6 times more frequent than the other individual pairs. For all SNP-pairs, minor allele frequencies ranged between 0.05 and 0.5 and LD ranged between an r^2^ of 0.5 and 0.9. In each individual, the genotype at the first SNP was generated conditional on the assumed allele frequencies, penetrance model, and the individual’s affection status. Then, pairs of haplotypes were generated by simulating alleles at the second SNP conditional on the allele at the first SNP, the assumed value of LD, and the penetrance model. Risk/null pairs had a relative risk range between 1.05 and 1.3 for the risk SNP, with the range selected to reflect typical psychiatric GWAS results. Inverse-risk/null pairs had a relative risk range between the reciprocal of 1.3 and the reciprocal of 1.05 for the inverse-risk SNP. For risk/resilience pairs, the base relative risk range for risk SNPs was 1.05 to 1.3 while the base relative risk for resilience SNPs ranged between the reciprocal of 1.3 and the reciprocal of 1.05.

When looking at SNP effect size individually, as is commonly the case in GWAS, the opposing effects of risk and resilience SNPs in LD make the odds ratios of each SNP appear smaller than the true effect when considering interactions. In real data, it is possible that SNPs with risk/resilience interactions are still among the top results in GWAS, which would mean that those SNPs have true effect sizes that are higher than those estimated from the GWAS. Therefore, to allow for correlated SNPs with opposing effects reducing the apparent effect sizes, we multiplied the base relative risk range of SNPs in risk/resilience pairs by a value representing 2 times the LD r^2^ value between the SNPs. We chose this value to keep the individual odds ratios of SNPs in the risk/resilience pairs closer to those of the other pair types. This allows the risk/resilience pairs to better reflect those we would expect to include and detect in real data analyses, since we use individual SNP GWAS results to determine inclusion in neural network models. The resulting odds ratios of the risk SNPs in risk/resilience pairs were still significantly less than those in risk/null pairs and the odds ratios of the resilience SNPs were significantly greater from the inverse-risk SNPs in the inverse-risk/null pairs (Supplementary Table 6 and 7). This indicates that the effect sizes in risk/resilience pairs were weaker on average than those in the other pair types. Supplementary Figure 3 visualizes the first ten SNP-pairs for each pair type and demonstrates the induced reduction in apparent effect size in risk/resilience pairs.

Based on the LD, relative risk, and allele frequencies, a value of 0, 1, or 2 was generated for each SNP, reflecting the number of minor alleles generated for that SNP for each individual. For each SNP, we calculated log-additive risk for each individual by estimating allelic SNP effects using a logistic regression model predicting case/control status and multiplying the number of alleles by the allelic effect. For each simulated pair, we calculated the total risk, defined here as the risk that includes both risk and resilience effects, using the expressions in Supplementary Table 8, dependent on the values of both SNPs in the pair. We used the expressions in Supplementary Table 8 to simulate multiplicative effects of resilience SNPs that multiplicatively counteract the effects of risk SNPs. For both total risk and traditional log-additive risk, we summed scores across all SNPs generated for each individual to create polygenic risk scores. We continued randomly generating SNP-pairs until the top 5% of total risk scores in the set of control simulations no longer completely overlapped with the bottom 5% of total risk scores in the case simulations.
